# Microfluidics-Enabled Digital Isothermal Cas13a Assay

**DOI:** 10.1101/2021.08.18.21262201

**Authors:** Frank X. Liu, Johnson Q. Cui, Hojeong Park, Ka Wai Chan, Tyler Leung, Ben Zhong Tang, Shuhuai Yao

**Affiliations:** Department of Mechanical and Aerospace Engineering, The Hong Kong University of Science and Technology, Clear Water Bay, Kowloon, Hong Kong; Department of Chemical and Biological Engineering, The Hong Kong University of Science and Technology, Clear Water Bay, Kowloon, Hong Kong; Department of Chemistry, Hong Kong Branch of Chinese National Engineering Research Center for Tissue Restoration and Reconstruction, The Hong Kong University of Science and Technology, Clear Water Bay, Kowloon, Hong Kong; DiagCor Life Science Limited, Kowloon Bay, Kowloon, Hong Kong; Shenzhen Institute of Molecular Aggregate Science and Engineering, School of Science and Engineering, The Chinese University of Hong Kong, Shenzhen,2001 Longxiang Boulevard, Longgang District, Shenzhen City, Guangdong 518172, China

## Abstract

The isothermal molecular diagnosis with CRISPR has attracted particular interest for the sensitive, specific detection of nucleic acids. However, most of the assays with Cas enzymes were performed in bulk assays using multistep approaches and hard to realize quantitative detection. Herein, we report Microfluidics-Enabled Digital Isothermal Cas13a Assay (MEDICA), a digital format of SHERLOCK with enhanced robustness and sensitivity. We first address the macromolecular crowding problems when combining the recombinase polymerase amplification (RPA) and Cas13a detection into a one-pot SHERLOCK assay. After the assay optimization, the enhanced one-pot SHERLOCK (E-SHERLOCK) achieves high robustness and 200-fold increased sensitivity. Leveraging droplet microfluidics, we streamline the E-SHERLOCK to eliminate undesired input targets caused by pre-amplification before partition, enabling background-free absolute quantification. From the real-time monitoring, MEDICA enables qualitative detection within 10 min and absolute quantification within 25 min. For the proof of concept, we applied MEDICA to quantify HPV 16 and 18 viral loads in 44 clinical samples, indicating perfect accordance with qPCR results. MEDICA highlights the CRISPR-based isothermal assays are promising for the next generation of point-of-care diagnostics.

## Introduction

Rapid, robust, and inexpensive diagnostics are crucial in defense against the spread of viral infections such as SARS-CoV, MERS-CoV, Ebola, Zika, avian, and swine influenza. The COVID -19 pandemic highlights the unprecedented demand for rapid and sensitive nucleic acid detection assays, which play a pivotal role in routine diagnosis, mechanistic and transmission studies, vaccine development, and therapeutics for COVID-19^1^. Quantitative polymerase chain reaction (qPCR) has long been regarded as a gold standard for viral infection diagnosis^2^. However, as a centralized laboratory-based testing paradigm for viral diagnostics, PCR assays depend on specifically designed TaqMan probes as well as precise thermal cycling, limiting their deployability for community tests.

Recently, clustered regularly interspaced short palindromic repeats (CRISPR) and CRISPR-associated (CRISPR-Cas) endonucleases are leveraged for nucleic acid detection through unique collateral trans-cleavage^3-10^. Programmable crRNA-guided CRISPR Cas12 and Cas13 can specifically recognize the target sequence to promote trans cleavage of surrounded single-stranded reporters^11, 12^. Combined with a separate step of isothermal recombinase polymerase amplification (RPA) preamplification, Cas13a based SHERLOCK (Specific High-sensitivity Enzymatic Reporter unlocking) and Cas12a based DETECTR (DNA endonuclease-targeted CRISPR trans reporter) achieved attomolar sensitivity and single nucleotide polymorphism (SNP) identification ability^13, 14^. Due to the strong collateral activity Cas13a^8, 15, 16^, the SHERLOCK assay demonstrated single-molecule detection ability when performing using a multistep process. However, the multistep schemes not only increase the risk of contamination and human errors but also impose difficulties and sample loss for the quantification of the target molecules. Even though SHERLOCK can perform at a one-pot scheme, the macromolecular crowding agent issue and enzymatic incompatibility between RPA and CRISPR reactions result in less robust and sensitive results compared to two-step reactions^17^. In addition, the one-pot reaction relies on the use of standard curves and endogenous controls to obtain relative quantification, which has been shown to have significant variations^18, 19^. In contrast to the standard curve calibration approach, absolute quantification with improved precision and accuracy has been achieved by digital PCR^20^ and digital loop-mediated isothermal amplification (LAMP)^21^. In digital quantification, the assay mixture is partitioned into a sufficient number of individual reactions so that either zero or one of the target nucleic acid molecules is being amplified in each partition, and the assay result is evaluated based on a simple positive-negative counting of the individual partitions based on the end-point fluorescence measurement^19^. Yet, as far as we know, there is no digital SHERLOCK assay available.

Different from PCR or LAMP that needs to be triggered by elevating the temperature, the isothermal RPA and some of the CRISPR type V and VI endonucleases can react at room temperature^22-24^ so that the reaction would start to amplify the targets before the sample partitioning into the nano-wells (e.g., QuantStudio 3D digital chips) or droplets using a simple flow-focusing system, which inevitably causes undesired overestimation due to pre-amplification^24-26^. To minimize the premature target amplification, one remedy is to prepare the reaction mixture on ice and quickly load it for partition^27^. Hence, unlike hot-start digital PCR and LAMP, the implementation of digital quantification of the near ambient temperature SHERLOCK needs a novel system for sample partitioning. Droplet microfluidics offers versatile yet controllable handling of samples and reagents in forming droplets, and thus can effectively avoid premature amplification before partitioning into droplets as well as facilitate combined amplification and trans-cleavage detection reactions in individual droplets.

In this work, we report Microfluidics-Enabled Digital Isothermal Cas13a Assay (MEDICA), the digital format of enhanced one-pot SHERLOCK (E-SHERLOCK). Digital quantification relies on a high signal-to-background ratio to convert the fluorescent signals of individual droplets to one or zero readouts, thus signal robustness and reproducibility are of great importance for digital quantification. However, when we conducted the one-pot SHERLOCK assay, the poor sensitivity and robustness of the reaction raised great concern. We identified the optimal reaction condition enzymatic incompatibility and addressed the macromolecular crowding agent issue of the all-in-one SHERLOCK assay, allowing robust detection at a single copy per microliter. We further applied E-SHERLOCK assay into droplet microfluidics for digital quantification. Leveraging the microfluidic design, MEDICA uses droplet compartmentation technology that effectively prevents premixing by separation of the magnesium initiator from the target-included master mix, which allows MEDICA to eliminate undesired premature amplification. Moreover, MEDICA showed an excellent signal-to-background ratio upon coordination of the reporter concentration with the collateral reporting, enabling near background-free absolute quantification. Benefit from the concentrated reaction within a picolitre droplet and the enhanced molecule mobility, MEDICA achieved qualitative detection within 10 min and quantitative detection within 25 min. Lastly, we applied MEDICA to detect both HPV 16 and HPV 18 for clinical validation. MEDICA successfully identified and quantified all viral HPV 16 and HPV 18 in clinical specimens, demonstrating excellent performance in line with qPCR results.

## Results

### Overview of MEDICA for viral HPV Detection

We established the workflow of MEDICA and demonstrated its application for the detection of viral Human Papillomavirus (HPV) 16 and 18. HPV is critical for identifying the risk of HPV-related cancers such as cervical cancer. Among all HPV types, HPV 16 and HPV 18 are accounted for the most precancerous lesions^28^. We designed two MEDICA sequences that include two sets of forward primers with overhanging T7 promoter, Cas13a crRNAs to recognize both HPV 16 and HPV 18 for specific and sensitive tests (Figure 1a). RPA is a magnesium acetate (MgOAc) initiation nucleic acid amplification near ambient temperature compared to hot start LAMP and PCR^29, 30^. Since RPA starts as soon as MgOAc is added even at room temperature^31, 32^, the premature RPA amplification caused by directly mixing MgOAc with the master mix before sample loading or partitioning would result in overestimated target input. To tackle this issue, we separate MgOAc and the target-included master mix before loading them into the droplet generator. Adopting a flow-flowing configuration with two sample inlets (Figure 1b), MgOAc and the target-included master mix are loaded separately into the device and co-encapsulated into microdroplets in fluorinated oil 7500 with 2.5% EA surfactant. Thus, RPA is only triggered by MgOAc after droplet formation. The droplet generator enables high throughput generation of monodisperse droplets of ∼30 μm in diameter at one kHz, which is more rapid and efficient than nano-well-based digital loading^33, 34^. By droplet partitioning, a single target DNA is confined in a small volume in which the signal can be amplified and concentrated for detection. The generated emulsions are then incubated at 37 °C before fluorescence imaging. This one-pot reaction contains extensible RPA amplification with T7 overhangs, RNA activator transcription by T7 RNA polymerase, and CRISPR Cas13a ternary collateral detection (Figure 1c). During the one-pot enzymatic activity, the unique adaptable reporting feature of Cas13a nuclease barely digests DNAs, leading to extremely high sensitivity. Our droplet-based MEDICA allows absolute counting of the target DNA molecules to achieve unprecedented accuracy.

**Figure 1.**
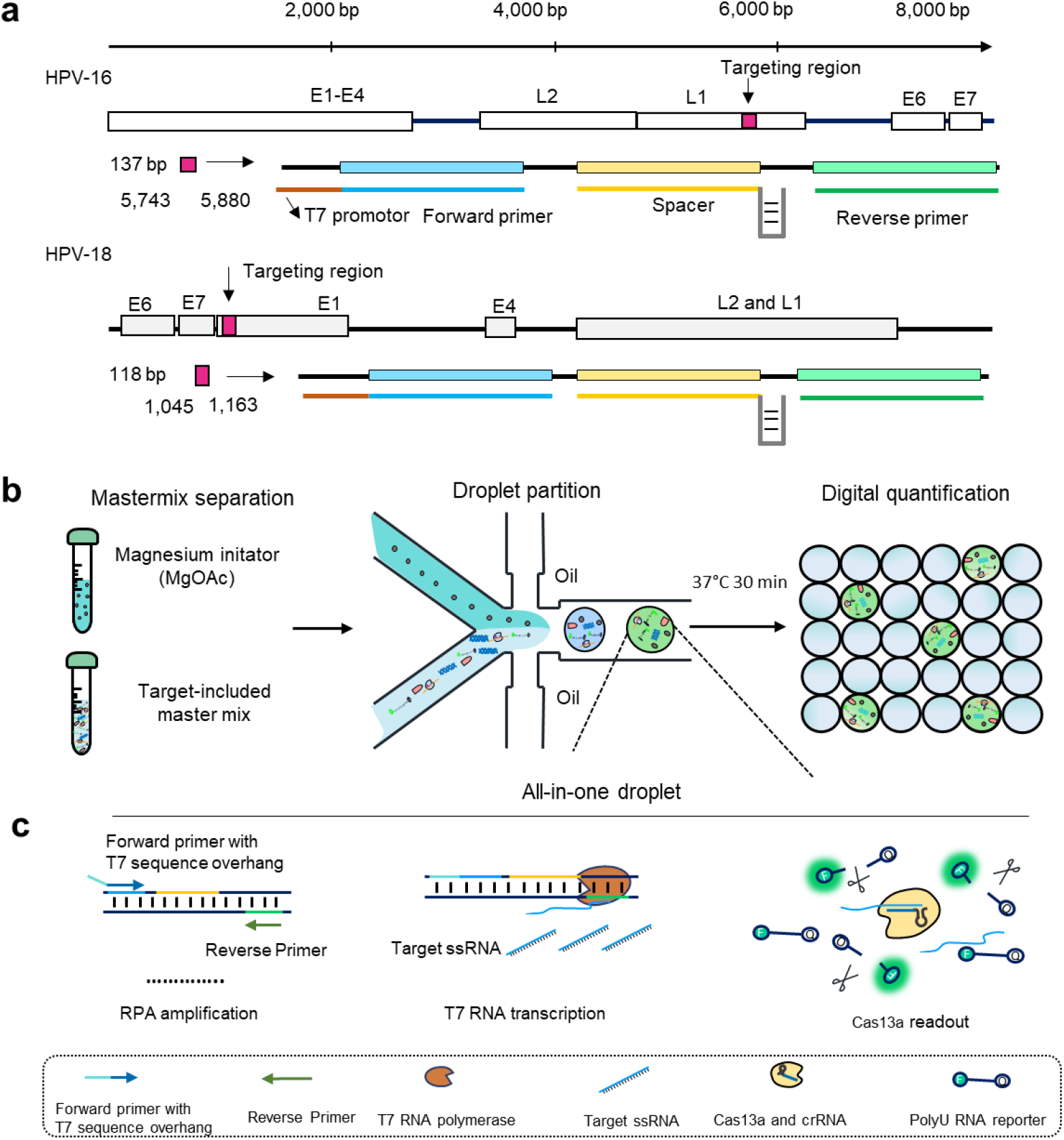
Overview of targeting sequences and MEDICA process. **a**, The genome map of both HPV 16 and HPV 18. The selected CRISPR detection regions of HPV 16 L1 and HPV 18 E1 are annotated with red lines in the genome, which covers the forward RPA primer with extended T7 region overhang, targeted protospacer, and reverse primer. The targeted HPV 16 region has 137 bp (from 5743 bp to 5880 bp) while the HPV 18 detection region has 118 bp (from 1045 bp to 1163 bp). Spacer represents the crRNA spacer. **b**, The workflow of the digital quantification process. A master mix with the extracted sample and magnesium initiator (MgOAc) are prepared separately and flow in the droplet generator as two aqueous phases. MgOAc triggers the reaction as soon as droplets are formed. After incubation at 37 °C for 30 min RPA and Cas13a reactions, the droplets are imaged by fluorescence microscopy and counted for quantification of the target molecule concentration. **c**, The all-in-one molecular reaction inside a droplet. With the initiation of MgOAc, RPA enzymes amplify the target sequence to generate numerous amplicons with a sequence extension of the T7 promoter region. Then T7 RNA polymerase transcripts target protospacer into RNAs to activate Cas13a for trans-cleavage reporting.

### Optimization of the one-pot SHERLOCK assay

Combining RPA with Cas13a in a one-pot reaction with high sensitivity and reproducibility is critical to ensure the accuracy and robustness of the quantification results. Therefore, we started with assay development which is a prerequisite to digital quantification. However, when we conducted the one-pot assay directly combining the reagents used in the two-step SHERLOCK ^35^, we found the results had dramatical fluctuation and thus compromised sensitivity compared to the two-step approaches (Figure 1a, supplementary S2). The digital quantification relies on the statistical calculation of the positive droplets that contain fluorescence signal higher than the threshold ^19^. The irreproducibility of the reactions in droplets would result in massive rain droplets whose signals are between the positive and negative controls, leading to a high error to absolute quantification^36^. Therefore, in circumventing these challenges, a compatible and robust one-pot SHERLOCK assay needs to be established in order to minimize the uncertainty in counting the positive droplets. These signal fluctuations and uncertainty are likely caused by the reaction component incompatibility and viscous condition^17, 37^.

Subsequently, we performed multiple rounds of assay optimization to identify the key factors of this reaction. First, we investigated the components of the reaction buffer including pHs, salts, and other additives. We found the 60 mM pH 7.5 Tris-HCl buffer with 80 mM KCl yielded the highest signal, much better than the HEPES buffer used for SHERLOCK detection (Figure 2b and supplementary S3a). The commonly used RPA mixture is highly viscous due to the presence of a high molecular crowding agent to enhance the catalytic activity and increase specificity as well as sensitivity^29^. Macromolecular crowding increases the binding of polymerase to DNA and reduces the thermodynamic activity, resulting in localized reagent depletion^38^. Thus, additional components in low concentration are difficult to be distributed homogeneously. We then explored the influence caused by PEG 8000. The reaction was activated with the presence of PEG 8000 (Figure 2c), however, as the concentration of PEG 8000 increased over 5 %, the fluorescence signal of the one-pot reaction fluctuated dramatically. This result indicated that the collateral activity was affected by the viscosity of the reagent as the biomolecules of the Cas13a system are restricted in RNA activator transcription, Cas13a ternary trans reporting. Compared to the hot-start PCR assisted by PEG, the molecule diffusion is enhanced with the high temperature where the non-concentration components are distributed homogenously. Hence, we chose 2.5 % PEG 8000 to minimize the signal fluctuation caused by nonhomogeneous molecule distribution while maintaining robust detection performance. Moreover, we added 0.05 % tween 20, a surfactant commonly used in droplet microfluidics^39^, to reduce the viscosity even further (Supplementary S3c). We compared the viscosity of the final enhanced reaction buffer (e-reaction) with other PEG-based reaction buffers (Figure 2d). The optimal one-pot reaction buffer is less viscous than most of other PEG-based reaction buffers. These improvements decrease the viscosity of the reaction buffer, in another word, the molecule diffusion ability was increased at a lower temperature. To further enhance the detection ability, we optimized the primer concentration for this one-pot reaction (Supplementary S4a). 120 nM and 360 nM primers resulted in a strong fluorescence signal and 360 nM primer performed better at 10 cp/µL concentration (Supplementary S4b). Besides, we optimized all the other related components including crRNA to Cas13a ratio, T7 RNA (Supplementary S4c and d).

**Figure 2.**
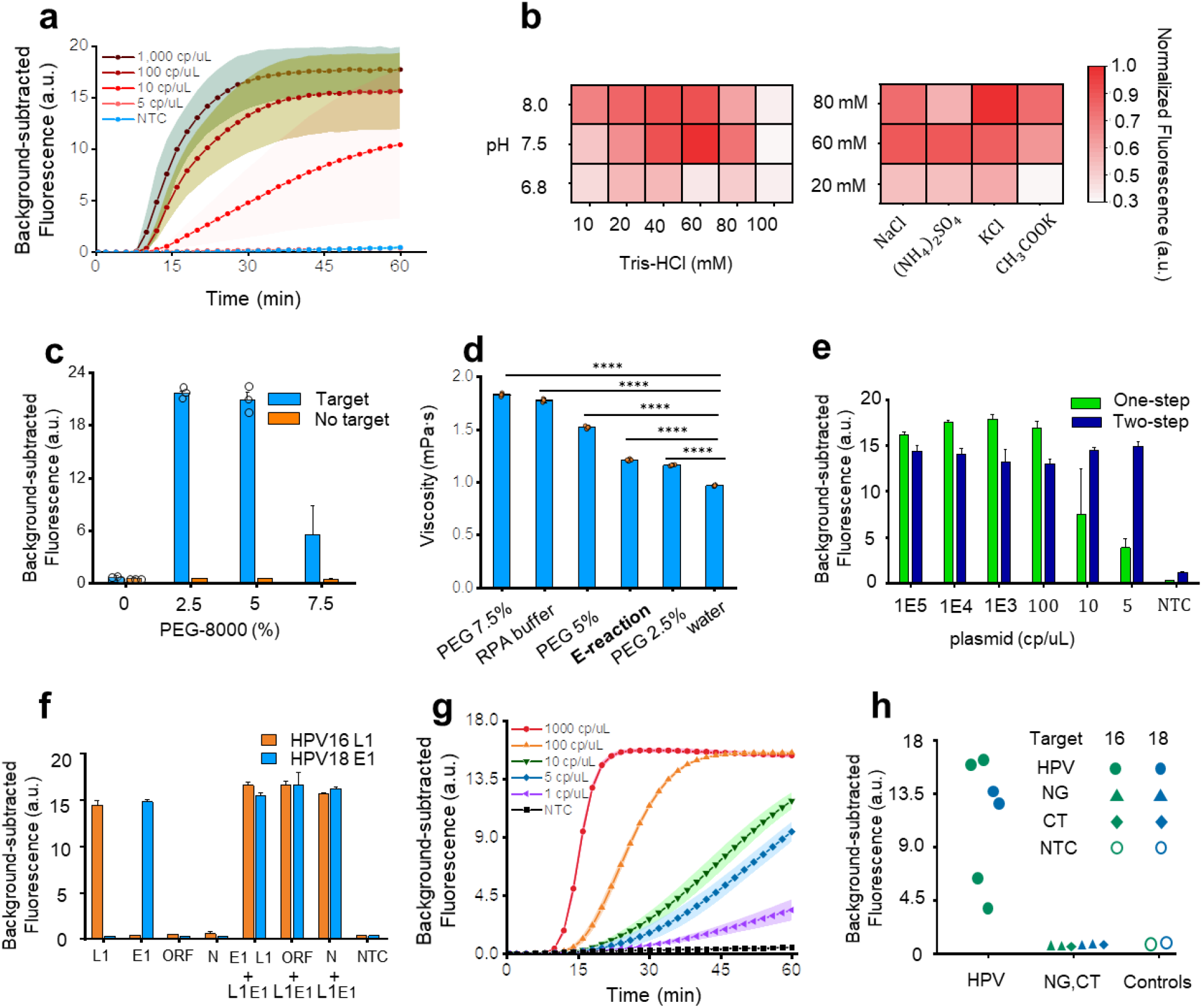
Construction of E-SHERLOCK. **a**, The fluorescence signals show a wide range of variations using the initial one-pot SHERLOCK in bulk for detection of synthetic HPV 16 plasmid from 1000 cp/μL to 5 cp/μL. **b**, The pH and salt optimization for one-pot SHERLOCK. Tris-HCl of pH 6.8, 7.5, 8, and different concentrations was compared. Sodium chloride (NaCl), amino sulfate ((NH_4_)_2_SO_4_), potassium chloride (KCl), and potassium acetate (CH_3_COOK) of different concentrations were compared. **c**, PEG 8000 of 0, 2.5, 5, and 7.5% was compared. **d**, The viscosity comparison of different reaction conditions using different concentrations of PEG8000, the e-reaction buffer represents the optimal reaction buffer. **e**, The comparison of two-step and the optimized one-pot SHERLOCK after 1 h using HVP 16 plasmid. **f**, The specificity of one-step SHERLOCK with or without spiked plasmid targets. ORF and N indicate the SARS-CoV-2 ORF gene and N gene, respectively. **g**, The real-time fluorescence of the optimized one-pot assay for detection of a serial dilution of HPV 16 plasmid from 1000 cp/μL to 1 cp/μL. **h**, The clinical validation of E-SHERLOCK. HPV 16, HPV 18, NG, and CT samples were tested to verify the one-pot SHERLOCK performance. The green color group represents the HPV 16 detection while the blue group shows the HPV 18 diagnosis. For **a** and **g**, the filled area represents the error bar (s.d.) of each group. For **c-f**, the center is the mean of 3 technical replicates, while the error bar indicates the s.d. For **d**, the two-tailed student’s t-test; ****P<0.0001. For **b** and **c**, the concentration of plasmid HPV 16 is 10 cp/uL. The NTC represents the non-template control.

After these major optimizations, we compared our E-SHERLOCK with the standard two-step SHERLOCK to test the sensitivity and specificity. The optimized assay indicated consistent detection ability (Figure 2e). Then we conducted the specificity of this assay to detect the HPV16 L1 gene, HPV18 E1 gene. Four synthetic plasmids containing HPV 16 L1 gene, HPV 18 E1 gene, SAR-CoV-2 ORF gene, and SAR-CoV-2 N gene, were applied. We spiked the targeted HPV 16 into the unrelated samples to compare the performance inside the complicated matrix (Figure 2f). E-SHERLOCK demonstrated high specificity with spiked samples. Lastly, we evaluated the sensitivity of the E-SHERLOCK. It achieved 1 cp/µL in a bulk solution (Figure 2g and Supplementary S7). The theoretical limit of detection (LOD) is 0.06 cp/uL (Supplementary S6). Compared to the initial one-pot assay, the optimized assay has improved both robustness and sensitivity of the assay (Supplementary S5). The optimized one-pot SHERLOCK increased a maximum of 76-fold of stability, and 200 times of sensitivity (Supplementary S5 and S6). We sought to verify the clinical performance of our enhanced one-pot SHERLOCK by conducting a clinical validation to detect both HPV 16 and HPV 18 (Figure 2h). Among other sexually transmitted diseases (STD) including Neisseria gonorrhoeae (NG) and Chlamydia trachomatis (CT), the one-pot SHERLOCK identified all clinical samples within 30 min, which is in 100% agreement with qPCR results (Table 1).

### Real-time and end-point quantification with MEDICA

The E-SHERLOCK achieved high sensitivity and robust detection ability. However, the reaction time (30-40 min) compared to the typical RPA reaction (20-30 min) is over-extended. Droplet digital methods can increase the localized reaction concentration thus shortening the reaction time. Additionally, the droplet digital technique offers more precise quantification results compared to real-time quantitative detection. Therefore, we applied droplet microfluidics to covert our optimal assay into digital format.

The droplet digital assay contains three steps, droplet partition, droplet incubation, and digital visualization. To realize the MgOAc and target-included master mix separation and mix, we separated the two phases and let them flow into a two-inlet droplet generator (Supplementary S8 a). Due to the different viscosity of the two phases, the pressures applied were adjusted to achieve an equal volume flow rate for mixing in droplets. The mixture of MgOAc and Target-included master mix was encapsulated into picoliter-sized water in oil droplets (∼30 μm) (Supplementary S9). The resulting monodisperse emulsion droplets were incubated at 37 °C. After incubation, droplets were imaged by fluorescence microscopy and analyzed for the target concentration quantification.

We firstly evaluated the total reaction time of MEDICA. A customized stage was developed to capture the images of *in-situ* MEDICA (Supplementary S11). In the real-time MEDICA, we were able to detect significant fluorescence signals within 10 min when significant numbers of droplets are present (Figure 3a). We found the positive droplet ratio increased dramatically in 10-15 min and reached a plateau after 20 min (Figure 3b). In terms of the total fluorescence intensity, it increased until 25 min when the signals in droplets became saturated and where the amplification time (Cq) is between 5 to 10 min (Figure 3c). This indicates that the real-time MEDICA only took around 5-10 min for detection which is more rapid than the corresponding assay in bulk solution. We postulated that this is because the RPA and CRISPR reactions are accelerated within a picoliter confined compartment, and the signals generated are concentrated to facilitate the detection. The enhanced mobility of molecules in a less viscous fluid in droplets also contributes to the mixing of the reagents and therefore shortens the overall reaction time. Similar results have been reported in which the reaction in droplets is accelerated^40, 41^.

**Figure 3.**
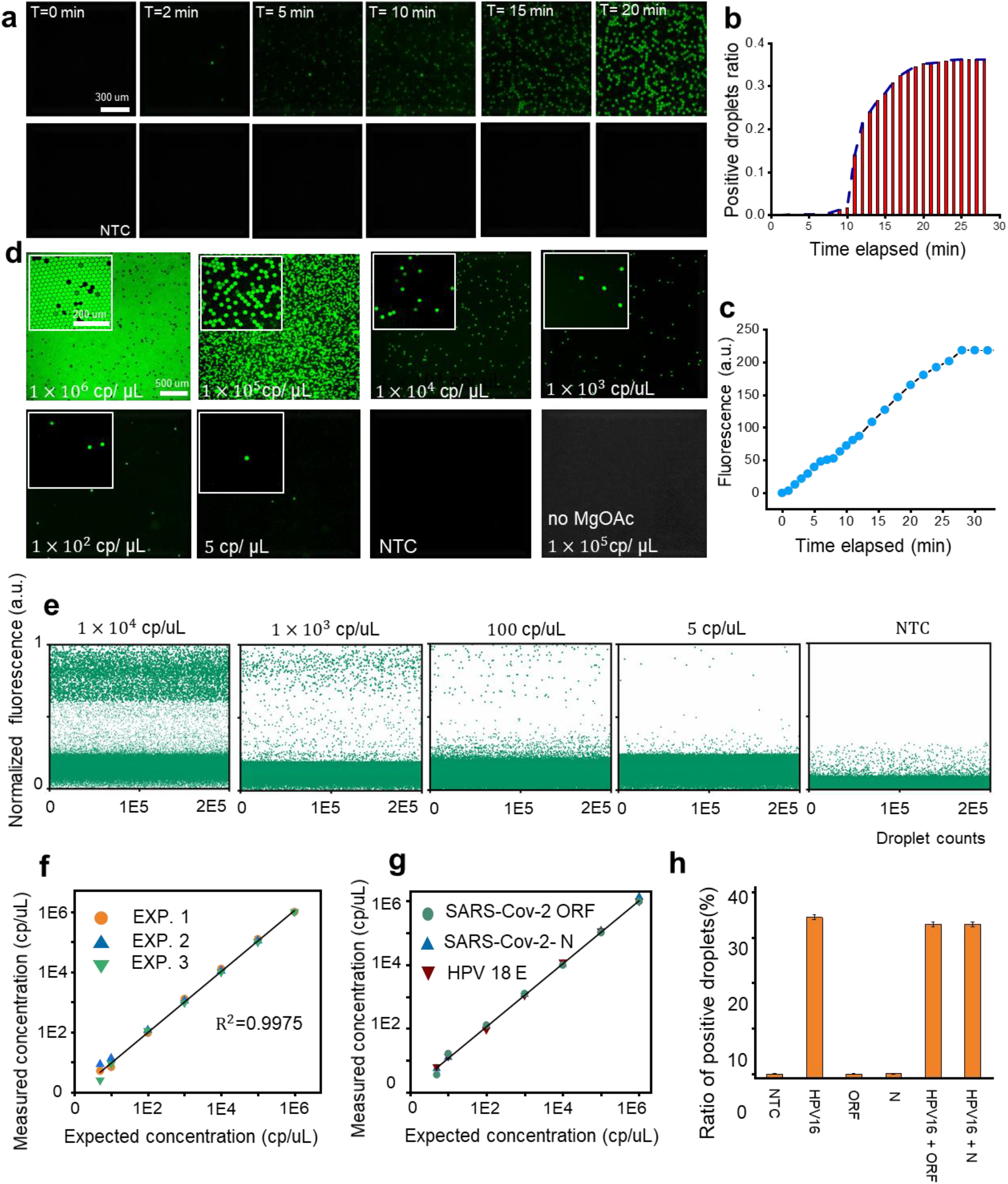
Real-time and end-point MEDICA. **a**, The fluorescence images of the real-time MEDICA (0-20 min) for quantification of 100,000 cp/uL HPV 16 plasmid. **b**, The percentage of positive droplet ratio along the reaction time. 10 min was the turnover point for qualitative detection, and the ratio was saturated around 25 min. **c**, The average intensity of all droplets over the reaction time. The highest fluorescence was yielded around 25 min. **d**, The fluorescence images of the end-point MEDICA using a serial dilution of the sample ranging from 1 × 10^6^ cp/µL to NTC. The extra control group was 1 × 10^5^ cp/µL with the absence of the MgOAc initiator. **e**, The statistical analysis of the end-point results. **f**, The measured concentration is in excellent agreement with the expected concentration. EXP1, EXP 2, and EXP 3 represent the three technical replicated experiments. **g**, Quantification of the SARS-CoV-2 ORF gene, SARS-CoV-2 N gene, and HPV 18 N gene by MEDICA. **h**, Specificity test of MEDICA to target HPV 16 with or without spiked into ORF and N gene groups. The concentration was 1 × 10^5^ cp/µL. Error bars represent the 95 % confidence interval.

Subsequently, we calibrated MEDICA by end-point quantification using serially diluted samples of synthetic HPV 16 plasmid. The results are shown in figure 3d. The absence of the MgOAc group emitted even less fluorescence on average than the non-template control (NTC) group, which further indicates that the reaction was completely suspended without the sole magnesium co-factor. The total and positive droplets were counted using ImageJ and customized Matlab code (Supplementary S10). Figure 3e shows the fluorescence intensity from the NTC group, which was below the threshold. The highest intensity of the recorded droplets from the NTC group is less than 0.5 after normalization, while all the positive droplets were selected over 0.6. Thus, MEDICA enables near background-free digital quantification. By tailoring the reporter concentration, the signal-to-background was steadily enhanced by increasing the reporting concentration (Supplementary S12), which can be attributed to the unique collateral cleavage of Cas13a. We obtained a linear correlation between the theoretical concentration and the measured concentration (Figure 3f). We were able to quantify as low as 6 copies in total 20 μL volume. Therefore, MEDICA demonstrated the single-molecule detection ability and high precision quantification from 0.3 cp/μL to 1,000,000 cp/μL. We further tested the robustness of MEDICA to quantify HPV 18 E1, SARS-CoV-2 ORF, and N genes. We observed the equivalent linear correlation of all the groups (Figure 3g), which further indicated the versatility of MEDICA. Finally, we tested the specificity of MEDICA in multiple conditions with or without the spiked samples. Taking HPV 16 as the main viral target, MEDICA successfully differentiated all the targets with consistent results (Figure 3h).

### Clinical validation of MEDICA

Finally, we demonstrated the feasibility of MEDICA to detect clinical viral HPV 16 and 18 samples. We evaluated the absolute quantification results of MEDICA with qPCR. After obtaining the standard curves of qPCR for HPV 16 and HPV 18, we compared the performance of MEDICA and qPCR. Both qPCR and MEDICA differentiated all the viral clinical specimens of HPV 16 and HPV 18 (Figure 4b and 4c). The concentration variation of the two methods indicates the same tendency. Since the C_t_ (threshold cycle) values straightforwardly represent the raw target concentration, we correlated the correlations of measured concentration from MEDICA with the C_t_ values of qPCR to verify the consistency of their results. The overall correlations of the measured C_t_ values and the quantified concentrations from MEDICA (Figures 4d and e) follow the expected trend. However, at the lowest concentration of HPV 18, the difference between these two methods tended to diverge. The absolution quantification tended to decrease slower than the real-time method. We believe these differences can be explained by two reasons. Firstly, when the input copies are limited, the qPCR quantification is likely to have significant differences in quantification results compared to the digital method^19^. Secondly, the concentration of the clinical samples may change after several rounds of freeze-thaw, resulting in this difference. The clinical validation further confirms that MEDICA offers rapid, accurate, and direct quantification of nucleic acids from clinical specimens.

**Figure 4.**
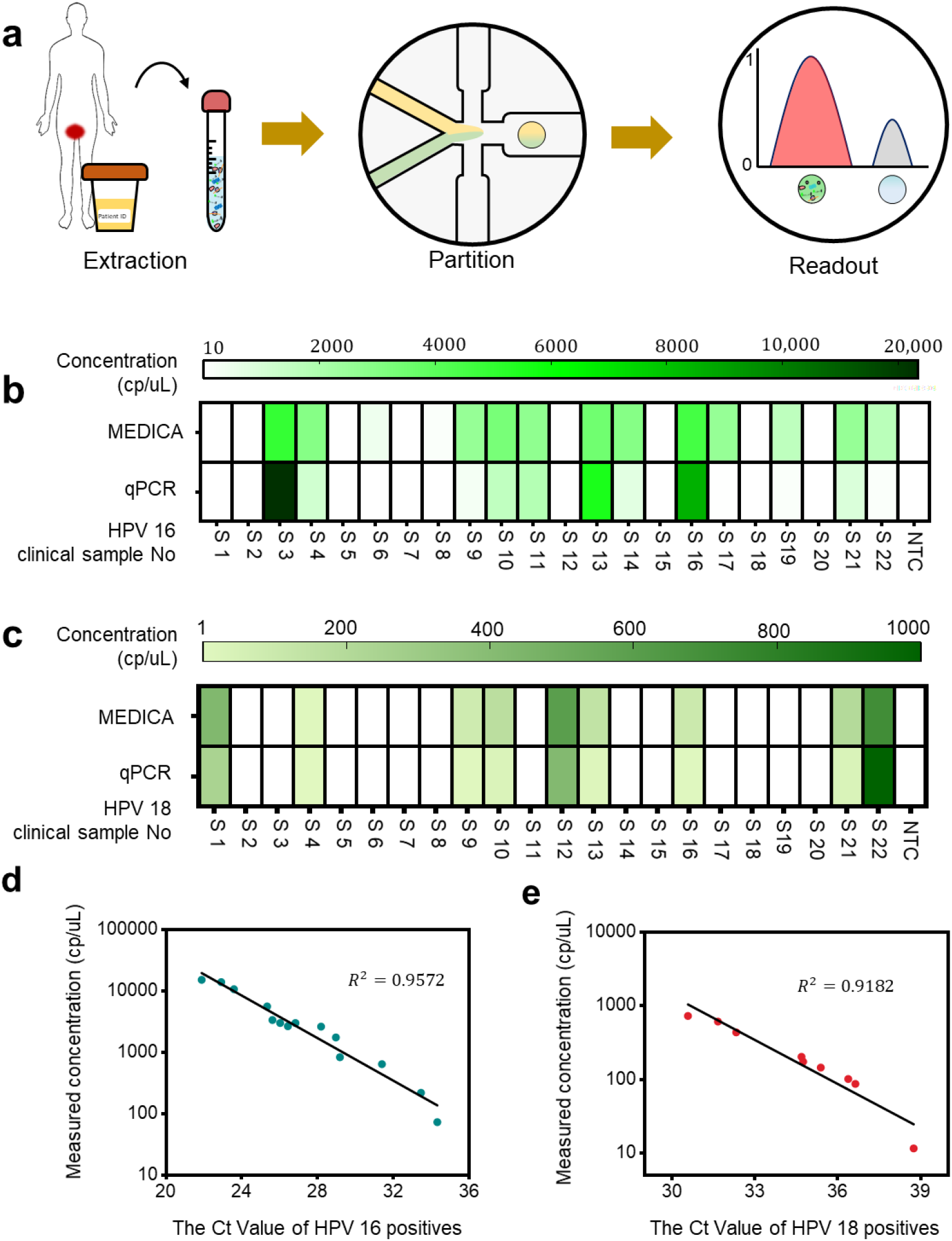
Clinical validation of MEDICA. **a**, Overview of MEDICA for viral HPV detection using clinical swab samples. **b**, Comparison of MEDICA quantification results with qPCR results for detection of HPV 16. MEDICA identified all clinical samples with the same tend of qPCR. **c**, Comparison between MEDICA quantification results and qPCR quantification of HPV 18. **D** Correlation between the HPV 16 concentrations measured by MEDICA and the real-time qPCR C_t_ values. **e** Correlation between the HPV 18 concentrations measured by MEDICA and the real-time qPCR C_t_ values.

## Discussion

Identification and quantification of trace amounts of nucleic acids are of great importance for public health and molecular biology study. Herein, we developed droplet digital quantification with a one-assay of RPA and Cas13a for viral nucleic acids (MEDICA). This microfluidic assay allows rapid and sensitive nucleic acid detection within 10 min and 25 min for absolute quantification. The enhanced one-pot reaches single copy per microliter detection around 35 min. When combined with droplet microfluidics, the droplet digital method confines the single-molecule within a single emulsion around picoliter volume to increase the localized concentration, shortening the reaction time. With simple ancillary equipment, MEDICA demonstrated a promising isothermal nucleic acid detection with Cas13a against rival thermal cycling PCR. The inborn collateral trans-cleavage of Cas13a is harnessed for signal amplification with the increase of molecule mobility, resulting in a uniform fluorescence readout.

Compared to previously reported one-pot hot-start CRISPR assays, our enhanced one-pot reaction required a lower reaction temperature, spotlighting the promising point of care diagnostics with simple requirements. Recently, the one-pot RPA with Cas12a and Cas13a was reported to attain high sensitivity through component optimization^4, 7^. However, these assays might be unable to achieve digital quantification due to irreproducible results by the influence of macromolecular crowding agents. This issue is originated from the RPA reaction, where the high molecular weight PEG hinders the movements of other molecules such as T7 polymerase, Cas13a nuclease, crRNA, and reporters, and thus affecting the trans-cleavage of reporters before or after CRISPR ternary activation. In PCR system, the high concentration of macromolecular crowding agent PEG is widely applied to enhance the sensitivity and specificity^42^. However, the high reaction temperature compared to the SHERLOCK test could increase the thermal diffusion of biomolecules, alleviating the inhomogeneous distribution. Therefore, at the near-ambient temperature reaction, the high concentration of PEG would inevitably affect the molecule diffusion. We reduced the concentration of the viscous PEG to define the trade-off between the RPA reaction and Cas13a readout for increased reproducibility. These improvements lead to the ultimate enhancement of trans-reporting of Cas13a. Thus, the optimized one-pot RPA and Cas13a reaction attain fast RPA reaction and powerful trans-reporting. Our optimal assay exhibits robust and reproducible detection ability at a single copy per microliter.

Digital assays near ambient temperature could easily induce inaccuracy during manual operations of sample preparation and partitioning. Hence, compartmentation technologies such as nano-wells and simple flow-focusing devices may lead to the overestimated detection results. We arranged the resuspension of the sole MgOAc initiator and master mix, enabling the reaction to be triggered after monodisperse emulsion. The microfluidic approach improves the accuracy and robustness of digital quantification. With the collateral enhanced one-pot assay, MEDICA achieved a uniform high signal-to-background ratio, avoiding rain droplets in discrimination of the positive and negative droplets. In addition, the picoliter scale reaction in droplets enhances the molecular mixing and collision in confined space, greatly promoting the efficiency of this assay. Because of its rapidity, sensitivity, specificity, and simple ancillary equipment, we expect that MEDICA holds great potential for system integration and miniaturization for point-of-care diagnostics.

## Materials and Methods

### Chemicals

Detailed information about reagents, including the commercial vendors and stock concentrations, all chemicals and sequences related to this work are listed in supplementary table 4 and table 5.

#### LwaCas13a protein extraction

LwaCas13a protein extraction was performed by following a previously published protocol with minor modifications^17^. *E. coli* Rosetta 2(DE3) pLysS transformed with the pC013 TwinStrep–SUMO–huLwaCas13a expression (Addgene plasmid #90097) and cells were cultured at 37 °C in 15 mL of Luria-Bertani (LB) medium containing 100 µg/mL ampicillin. Large-scale growth cultures were initiated by the addition of 5 mL of the starter cultures into 1 L of LB medium containing 100 µg/mL ampicillin and allowed to grow at 37 °C until the OD_600_ reached the range of 0.4 to 0.6. The cultures were transferred to 4 °C for 30 min to allow them to cool before the protein induction. The cultures were grown in a pre-chilled 16 °C biological shaker for 18 h with 0.1 mM IPTG. Cells were harvested, resuspended in 4X (wt/vol) supplemented lysis buffer composed of 20 mM Tris-HCl (pH 8.0), 0.5 M NaCl, 1 mM DTT, 2 cOmplete Ultra EDTA-free tablets, 100 mg of lysozyme, and 1 uL of benzonase to 60 mL of lysis buffer, and ruptured on ice by sonication (Q500, QSONICA). The lysate was centrifuged for 60 min at 10,000 rpm. at 4 °C. The supernatant was collected into a 50 mL falcon tube, and 0.5 mL of Strep-Tactin superflow Plus resin was added to the supernatant. The recombinant protein was bound to the resin for 3 h by gentle shaking at 4 °C. The resin-sample suspension was poured into 50-mL Bio-Rad glass Econo-Column equilibrated with cold lysis buffer. After flow-through was removed, the column was washed with 50 mL cold lysis buffer three times. 5 mL of SUMO protease cleavage solution composed of 5 mL of lysis buffer, 50 µL of SUMO protease, and 7.5 µL of NP-40, was added to the column, followed by and gentle shaking at 4 °C for 18 h. The cleavage solution was collected, and the column was washed three times with 5 mL of lysis buffer. The collected cleavage solution was dialyzed in 2 L of 1X PBS in a cellulose membrane dialysis tube for 18 h. The presence of recombinant was identified using SDS-PAGE gel analysis (Supplementary S1). Amicon Ultra-0.5 mL centrifugal filter was used to concentrate protein and re-diluted in protein storage buffer, which allows the protein for storage at -80 °C for up to 6 months.

#### Clinical extraction and qPCR validation

Clinical swab samples were provided by DiagCor Bioscience from clinical doctors in compliance with ethical and safety approval by Human Research Ethics Committee and Health, Safety and Environment Office of HKUST.

We used QIAamp DNA Mini Kit (Qiagen) to extract the viral RNAs according to the manufacturer’s protocol. The raw urine sample is 10 mL each. After extraction, samples were collected with a 1.5 mL tube and stored at -20 °C before use.

The qPCR was performed with a master mix of 20 μL including 10 μL of 2X Taq polymerase, 0.5 μL of 10 uM forward and reverse primers each, 0.5 μL of 10 μM probes, 5.5 μL of nuclease-free water, and 2 μL of target sample from the extraction. After gentle mixing, the two-step PCR was conducted with 30 s pre-heating followed by 45 cycles of 5 s at 95 °C (denaturation) and 30 s at 60 °C (annealing and extension). All the qPCR experiments were conducted with a Roche Lightcycler 480 II (Roche) at HKUST BioCRF.

#### One-step and two steps SHERLOCK

The one-step SHERLOCK was conducted with a 20 μL volume for each test. For experimental preparation, one lyophilized pellet was resuspended with 100 μL of final master mix with 10 μL of target and 5 μL of MgOAc (280 mM) deprived. The master mix was firstly prepared including 59 μL of rehydration buffer from the kit, 4 μL of LwaCas13a (1 μM), 4μL of rNTP mix (25 mM), 1 μL of Rnase inhibitor Murine (40 U/μL), 4.8 μL of each forward and reverse primers (10 μM), and 1.25 μL of Poly(U) reporter (10 μM). After reconstitution with one lyophilized pellet with gentle mixing, this mixture was aliquoted into 5 groups (17 μL). Then, 2 μL of target sample and 1μL of MgAOc (280 mM) were pipetted into accordingly with quick mixing before incubation. The sample was incubated with a Roche Lightcycler 480 II (Roche) at 37 °C. The fluorescence was measured every minute for one hour.

The two-step SHERLOCK is divided into two aspects: First, the RPA pre-amplification was triggered by adding MgOAc from the kit instructed with TwistAmp® Basic at 37 °C for 20 min. Second, 1 μL of the RPA product was added to 19 μL of Cas13a detection master mix consisting of 20 mM HEPES (pH 6.8), 9 mM MgCl_2_, 1 mM rNTP mix, 40 nM LwaCas13a, 1 U/μL RNase inhibitor murine, 0.125 U/μL T7 RNA polymerase, and 125 nM poly(U) RNA reporters. The fluorescence from the Cas13a trans-cleavage detection was then measured every 30 s using a Roche Lightcycler 480 II at 37 °C for 30 min.

#### Buffer construction and optimization for enhanced one-pot assay

The optimization of components of RPA and Cas13a reaction started with an initial reaction including 1X buffer containing 50 mM Tris-HCl (pH 8.0), 60 mM NaCl, 5% PEG 8000, and 2 mM DTT. The initial master mix was created with 100 nM Cas13a, 1 mM rNTP mix, 10 U of RNase inhibitor Murine, 100 nM crRNA, 5U of T7 RNA polymerase, 125 nM PolyU reporters, and 480 nM of each forward and reverse primers, and one lyophilized pellet from the basic RPA kit. In all experiments, the master mix without MgOAc was used to suspend the lyophilized pellet (one pellet to 100 μL final reaction volume). After reconstitution, this master mix was aliquoted accordingly, each group contains 17 μL of reconstituted master mix. Then, 2 μL of the synthetic sample or viral specimen was pipetted into this master mix with thorough mixing. Finally, 1μL of MgOAc (280 mM) from the kit was added to trigger this reaction.

Then optimization occurred iteratively with each reagent modification. We identified each component to yield the best signal then kept the optimal reagent tested previously when conducted the next optimization. We evaluated the performance of Tris-HCl with different concentrations (10, 20, 40, 60, 80, and 100 mM) as well as pH (6.8, 7.5, and 8.0). For the salt components, we tested NH_4_(SO_4_)_2_, NaCl, KCl, and CH_3_COOK at different concentrations (20, 60, and 80 mM), respectively. Then, we optimized the PEG-8000 concentration (0, 2.5, 5, and 7.5%). For the additives, we tested the Tween 20 (0, 0.05, 0.1, 0.2, and 0.5%) and DTT (0, 1, 4, 10, and 20 mM), respectively. After the buffer construction, we further optimized primer concentration (60, 120, 240, 360, and 480 nM), crRNA to Cas13a ratio (0, 0.5, 1, 2, 3, and 3.75), T7 polymerase (0, 0.1, 0.4, 0.8, and 2 U/μL) respectively. All the modulated reaction components are shown in figures and associated figure captions.

#### E-SHERLOCK in bulk

The final one-pot assay contains 1X reaction buffer (60 mM Tris-HCl pH 7.5, 80 mM KCl, 2.5% PEG-8000, 0.05% Tween 20 and 1 mM DTT), 40 nM Cas13a, 1 mM rNTP mix, 10 U Rnase inhibitor Murine, 40 nM crRNA, 0.8 U/μL T7 RNA polymerase, 125 nM Poly (U) reporters, and 360 nM each forward and reverse primers, and the enzymes from the lyophilized pellet (one pellet to 100 μL final master mix including the target). Once the master mix is created, it is used to resuspend the lyophilized pellets. The final reaction volume is 20 μL containing 17 μL of aliquoted master mix, 2 μL of target sample, and 1μL MgOAc (as the last component to start this reaction).

We further evaluated the performance of the sensitivity, specificity, and comparison of this one-pot reaction. All the experiments were conducted in real-time with the Roch Lightcycler 480 II with fluorescence measurement every minute at 37 °C for 1 h. For the specificity test, the spiked genes were firstly prepared at 1 × 10^5^ cp/µL and mixed mutually before experiments.

#### Droplet microfluidic device design and fabrication

The microfluidic device consists of Y-shaped two inlets for MgOAc and target-included master mix respectively, and two symmetric oil channels (fluorinated oil 7500 with 2.5 % EA surfactant) to form a flow-focusing droplet generator. The thickness of this device is 35 μm, and all the inlets are 40 μm until the flow-focusing junction with tiny modification. The aqueous phase channel of the junction is 30 μm while the oil channel is 25 μm. The outlet channel is 100 μm till the end.

The device was made of polydimethylsiloxane (PDMS, Sylgard 184 silicone elastomer, Dow Corning) using a soft lithography process. First, photoresist SU8 (3025) provided by NFF was deposited on a silicon wafer by spin coating. Then, the coated wafer was taken for micropatterning by employing photolithography (SUSS Mircotec MA6, Garching, Germany). Finally, the wafer was baked and developed (SU-8 developer; MicroChem, Westborough, MA, USA), forming a master for PDMS replica molding. To facilitate the molding of the PDMS device, Trichloro (1H, 1H, 2H, 2H-perfluorooctyl) silane 97% (Sigma-Aldrich, St. Louis, MO, USA) was used to coat the surface in a desiccator. PDMS mixture was prepared by mixing PDMS and solvent at a weight ratio of 10:1, which was poured on the silicon wafer. After baking at 80 °C for 1 hour, the PDMS layer was peeled off from the silicon master and then cut into an individual section. The final microfluidic chip was formed after bonding the PDMS to a glass substrate after oxygen plasma treatment. The microchannels were rendered hydrophobic after being treated with Aquapel, which is suitable for water-in-oil droplets formation. After binding, the microfluidic chip was firstly pre-treated with aquapel (PPG Industries) to yield hydrophobicity and followed by fluorinated Novec 7100 (3M) washing for 30 s. The microfluidic chip was incubated at a hotplate at 80 °C before use.

#### MEDICA quantification

The experiment was conducted with a pressure box (Everflow) to control the three phases (magnesium phase, master mix with target and oil phase). The total reaction volume is 20 μL containing two phases with equal volume. the 10 μL of magnesium phase contains 1X reaction buffer, 28 mM MgOAc. 10 μL of the other phase includes 1X enhanced reaction buffer, 80 nM Cas13a, 2mM rNTP, 16 U of Rnase inhibitor Murine, 80 nM crRNA, 1.6 U/μL T7 polymerase, 720 nM each forward and reverse primers, 1 μM poly(U) reporters, and 2μL of 1X target sample.

For end-point MEDICA, after 30 min incubation at 37 °C, droplets were pipetted into a 30 μm thick microchamber for fluorescence imaging. For the real-time MEDICA, firstly a heating system with a Peltier on microscopy was made with a PMMA frame. The temperature was calibrated to 37 °C. The sample was partitioned then collected on ice. Then, all the droplets were pipetted into a silicon chamber (30 μm) on ice. The inlet and outlet were sealed with optical adhesive film (Thermo Fisher). The time (0) image was recorded without heating. Then, the heating source was opened to record the fluorescence signal evolving with time. We used the synthetic SARS-CoV-2 ORF plasmid as an example to measure the real-time fluorescence change. The master mix was prepared with 1 × 10^5^ cp/µL target input, following the same protocol as MEDICA. The amplification time (Cq) for classifying samples as positive or negative was set to approximately 10 % of the average steady state fluorescence signal.

The droplet imaging system includes a LED light source with a 488 nm filter and a Nikon microscope. All the information (number and fluorescence intensity) was calculated by ImageJ (NIH). The detailed process of image analysis is listed in supplementary S10. According to the Poisson law, the average number of target DNA per droplet *λ* was calculated through the possibility of positive droplets (*p*_*postive*_). Let *M* to be the total number of droplets and *V*_*droplet*_ to be the average droplet volume. The molar concentration of the target *n*, can be

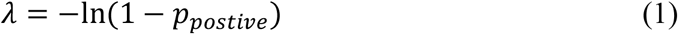

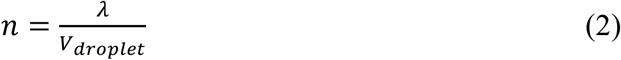

## Data Availability

all the data are available

## Acknowledgments

We would like to thank HKUST NFF for microfluidic chip fabrication, and HKUST BioCRF for the essential equipment supports. This work is supported by the University-Industry Collaboration Programme (Grant No. UIM/362) under Innovation and Technology Fund of Hong Kong.

## Author contributions

S.Y. and B.T. conceived and supervised the project. F.L., Q.C. and H.P designed the research. H.P purified the proteins and performed the verification. F.L. developed the assay and clinical PCR validation. F.L. and Q.C developed the microfluidic chip and conducted the droplet quantification experiments. Q.C developed the method for droplet analysis. K.C and T.L collected and purified all the clinical samples. F.L., Q.C., and H.P., analysed the data. F.L., Q.C., H.P., and S.Y. wrote the manuscript. All authors edited the manuscript.

