## Supplementary file for "Microfluidics-Enabled Digital Isothermal Cas13a Assay"

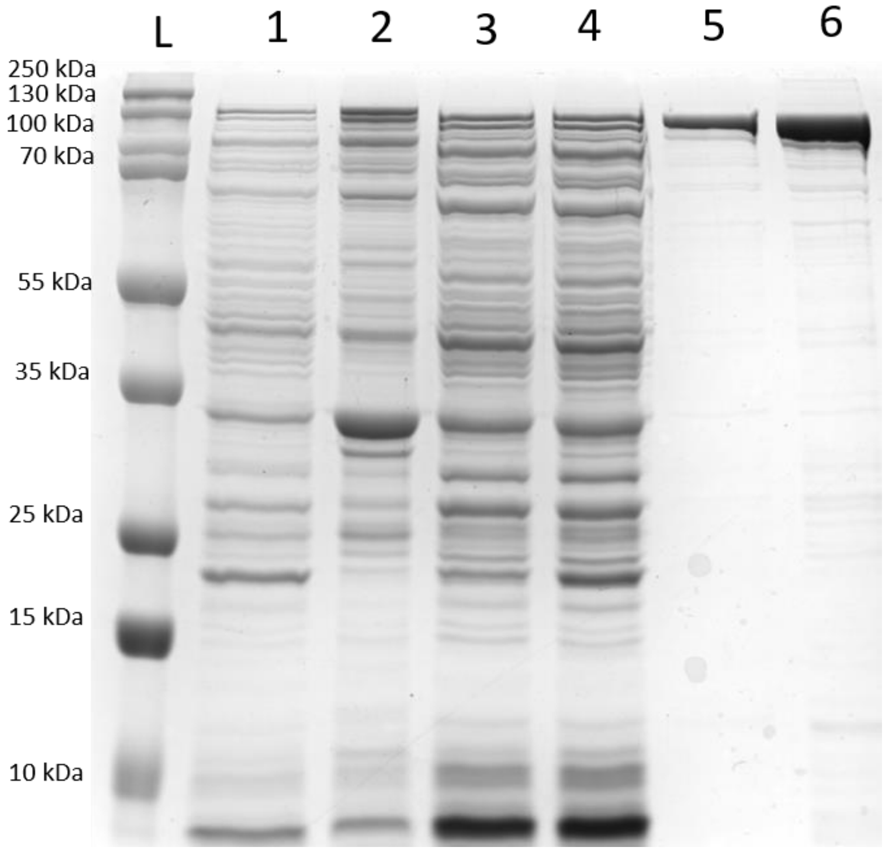

Figure S1. **Coomassie-stained SDS-PAGE gel of protein fractions**. Each line indicates L, ladder; 1, whole-cell lysate; 2, cleared cell lysate; 3, cell pellet after clearing of lysate; 4, flow-through following Strep-Tactin batch binding; 5, eluted fraction post SUMO protease cleavage; 6, concentrated sample after dialysis and ultracentrifugation. The purification protocol follows *(1)* with minor modification.

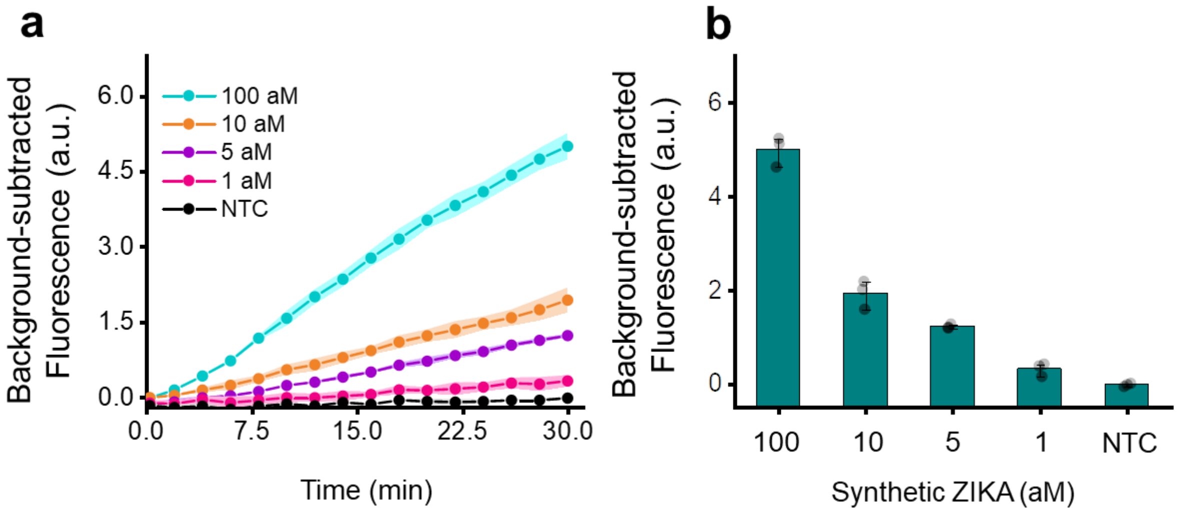

Figure S2. **Two-step SHERLOCK Test and Cas13a collateral reporting test**. **a,** The collateral detection of CRISPR Cas13a after 20 min RPA. The shaded area indicates the standard deviation (s.d.) of three repeated experiments. **b,** The endpoint fluorescence intensity of the two-step SHERLOCK for synthetic ZIKA detection following the protocol in *(1, 2)*, which exhibits the excellent sensitivity of SHERLOCK. The center equals the mean value of three replicates, while the error bar represents the s.d.

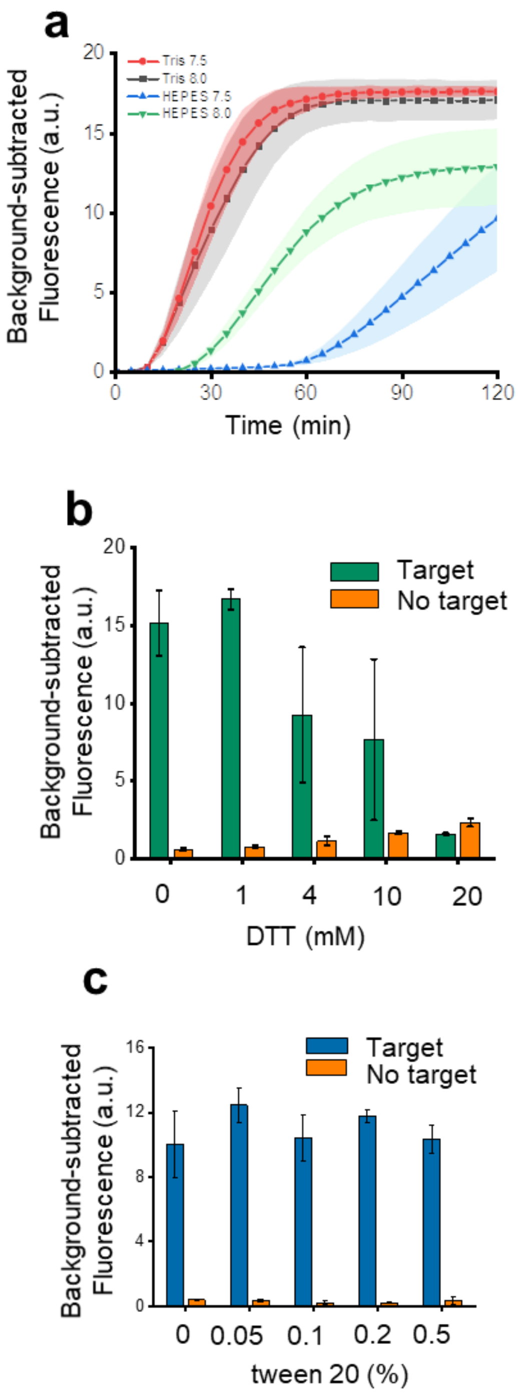

Figure S3. **The construction of one-pot SHERLOCK**. **a,** The performance comparison between Tris Buffer and HEPES buffer. **b,** Optimization of DTT for this reaction. c, Optimization of tween 20 surfactant. For **a,** the shaded area indicates the s.d. of three repeated experiments. For **b** and **c,** the center is the mean value of three replicates, while the error bar equals s.d.

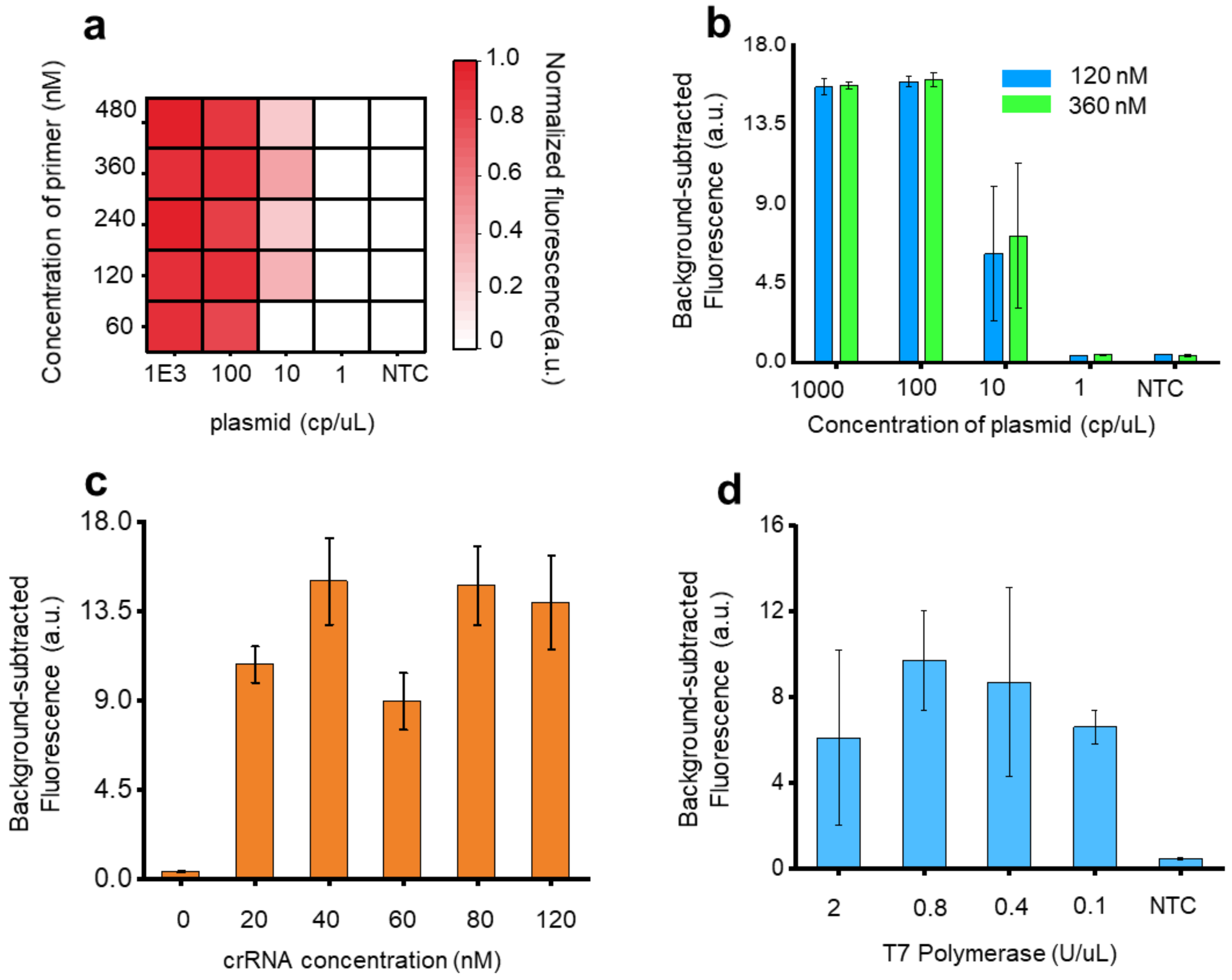

Figure S4. **Optimization of the reaction components of one-pot SHERLOCK**. **a and b**, Optimization of the primer concentration using HPV 16 plasmid. **c,** Optimization of the crRNA concentration using 10 cp/uL HPV 16 plasmid. **d,** Optimization of the T7 polymerase concentration. For **a,b,d**, incubation time was 1 h. For **c** and **d**, the HPV 16 plasmid concentration was 10 cp/uL. For **b, c** and **d,** the center equals mean value of three replicates, and the error bar is the s.d.

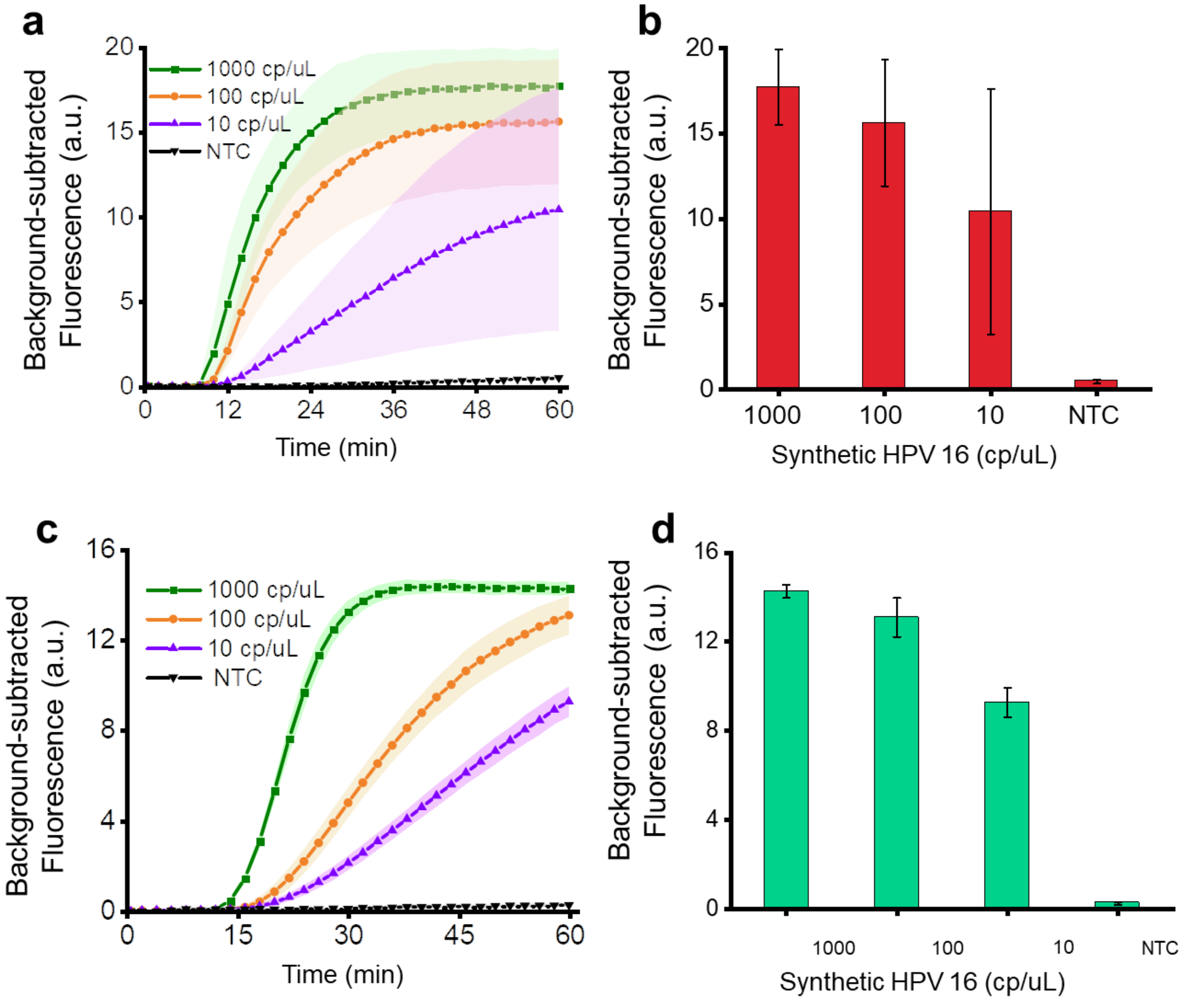

Figure S5. **The comparison between initial and enhanced one-pot SHERLOCK**. **a** and **c**, The time-course performance of the one-pot SHERLOCK at different concentrations before and after optimization. **b** and **d**, End-point comparison after 1 h. The signal fluctuation was dramatically reduced after optimization. The maximum decrease of the fluctuation is 76-fold after optimization. For **a** and **c**, the filled curve represents the error bar (s.d). For b and d, the center equals the mean of technical replicates, while error bar represents the s.d.

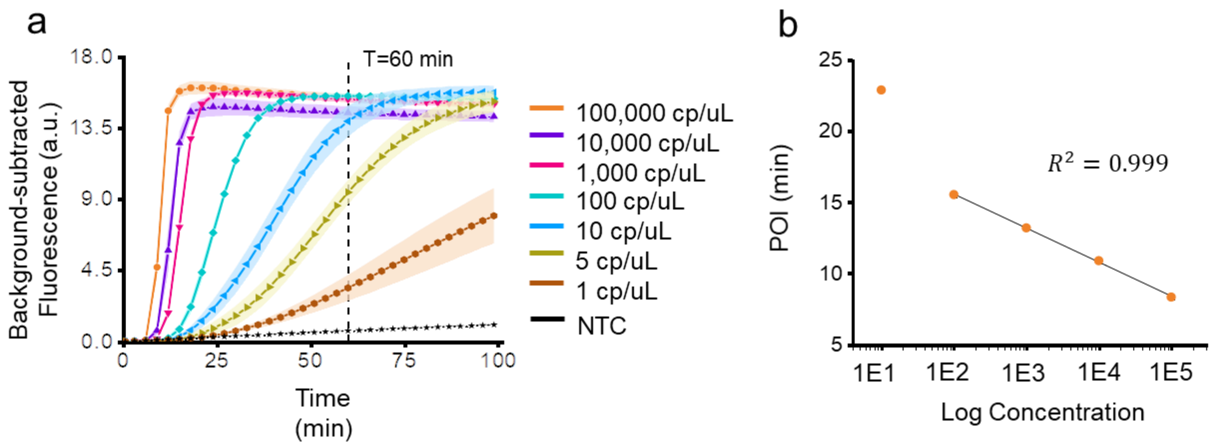

Figure S6. **Sensitivity evaluation of the enhanced one-pot SHERLOCK**. **a,** The assay performance using a serial dilution from a 1 cp/uL to 1,000,000 cp/uL. The theoretical limit of detection (LOD) calculation using 3σ (s.d. of blank) *(3)*. The calculated LOD is 0.06 cp/uL. **b,** The standard curve between the log concentration and point of inflection (POI, the time corresponding to the maximum slope in the fluorescence intensity curve)*(4, 5)* for real-time quantification between 100 cp/uL to 100,000 cp/uL.

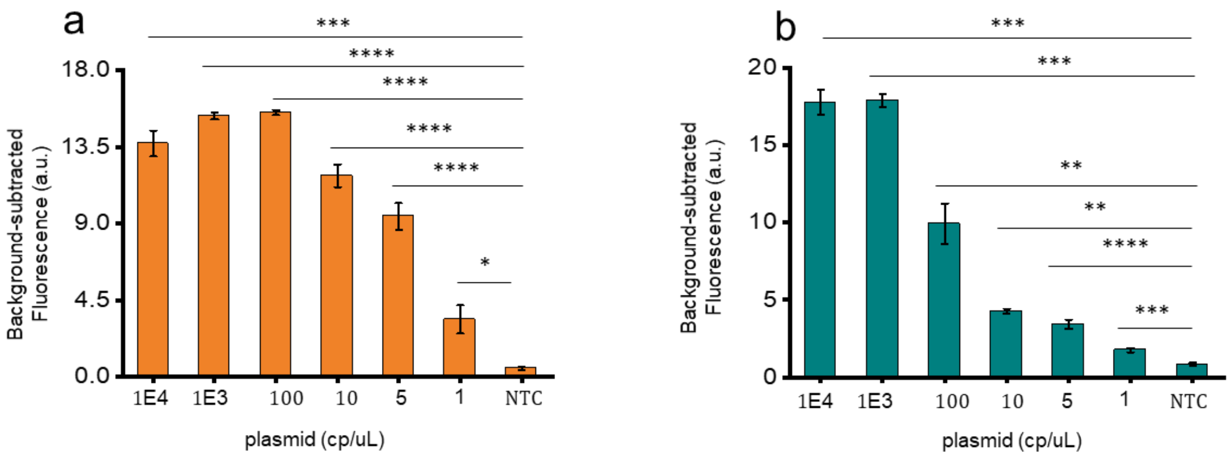

Figure S7. **Sensitivity test for both HPV 16 and HPV 18** . **a**, Sensitivity test for HPV 16. **b,** Sensitivity test for HPV 18. The center equals the mean value of three replicates, while the error bar represents the s.d. For the two-tailed Student's t-test, *P<0.05; **P<0.01; ***P<0.001; ****P<0.0001.

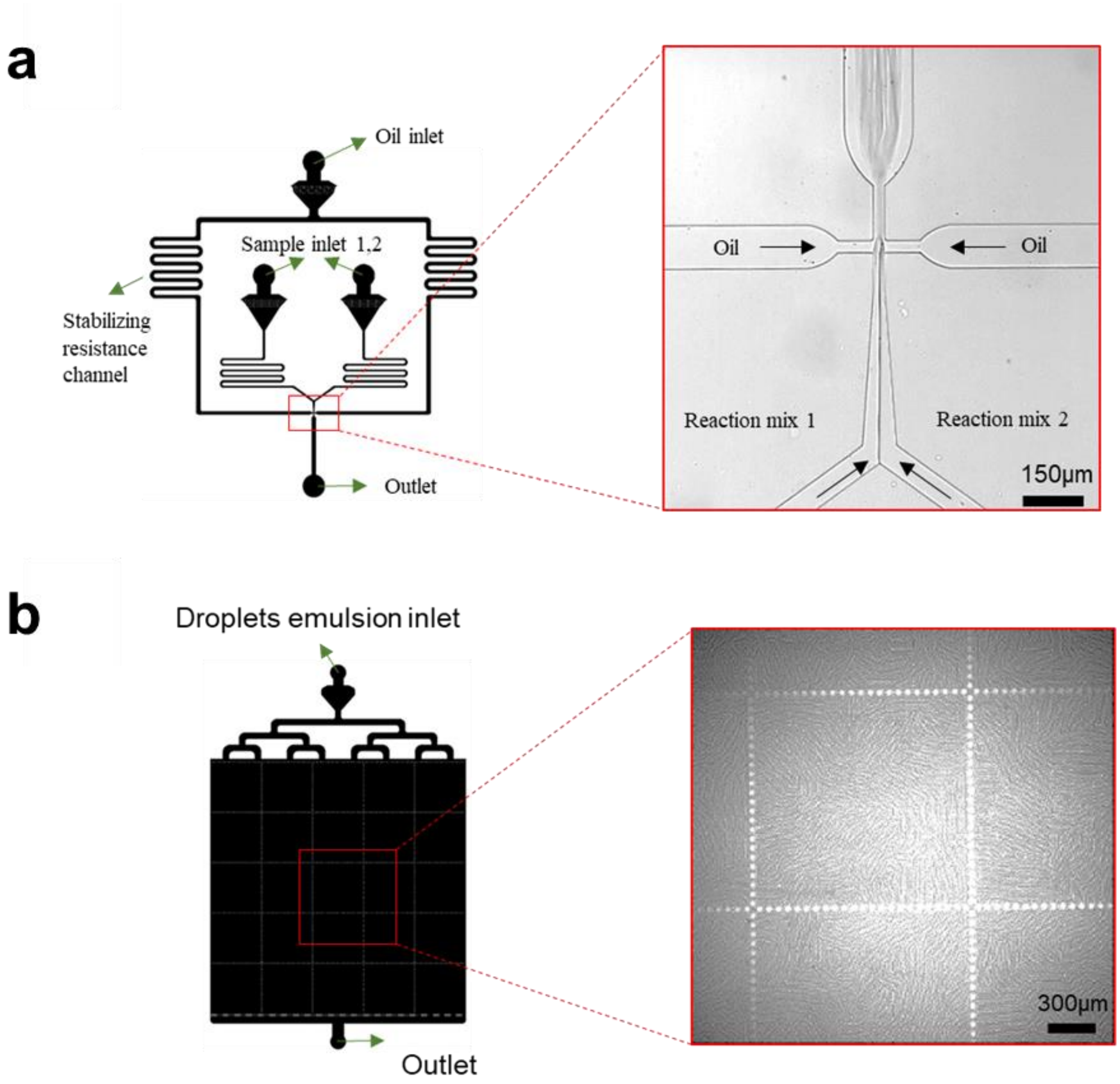

Figure S8. **Microfluidic chip design and layout**. **a,** The design of the droplet generation chip, the two-sample inlets separate the reaction initiator from other reaction master mix; microscope image of the droplet generation junction. **b,** The design of the droplet observation chamber chip, droplets are confined by micropillars for better imaging; microscope image of the droplets view.

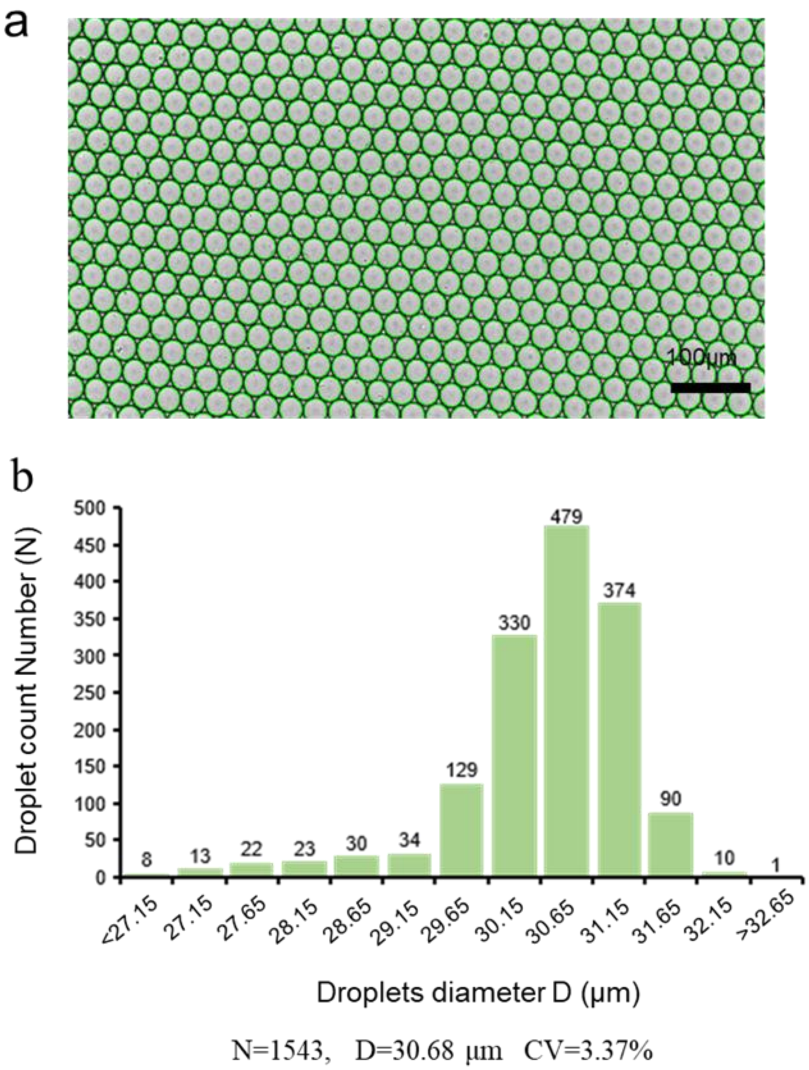

Figure S9. **Characterization of droplet size and size distribution**. **a, While field of droplets. b,** Droplet diameter distribution. The droplet diameter is measured by self-customized Matlab code. The average diameter is 30.68 $\mu$m and CV is 3.37%.

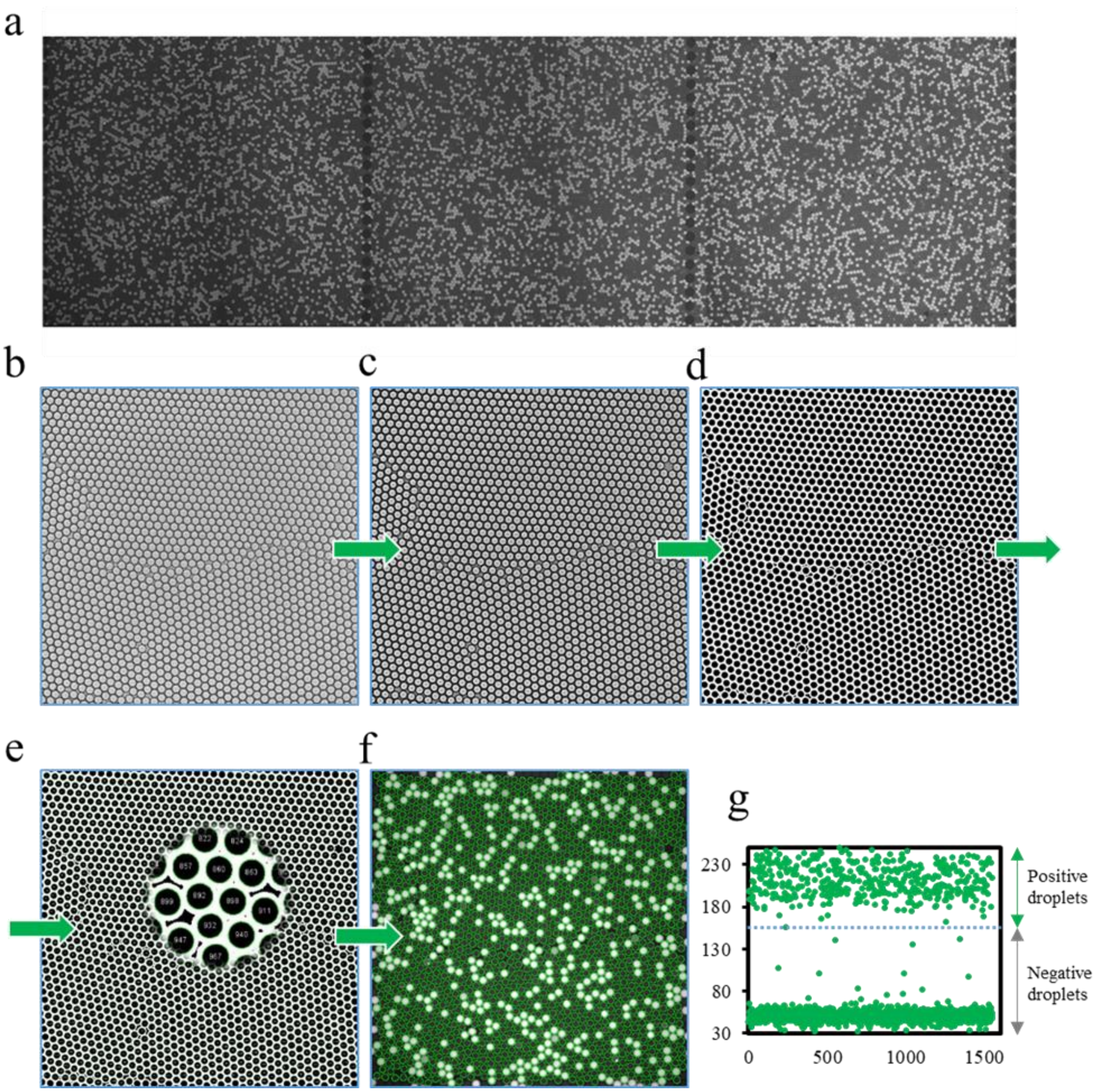

Figure S10. **Droplet fluorescence analysis**. The image processing is conducted in imageJ with a self-designed algorithm. **a,** An extended view of the droplet fluorescent image. **b,** Brightfield view of droplets. **c,** Image is defocused to show clear boundaries. **d,** Droplets are individually segmented by their boundaries. **e,** Each droplet is being identified and labeled, droplets location information is extracted as region of interest (ROI), in this process, thresholds regarding droplet area and circularity are set properly to avoid the selection of undesired merged droplets, non-specific dust and damaged droplets. **f,** ROI is used to mapping the droplets location in the corresponding fluorescence image, and the fluorescence intensity of each ROI is measured and exported. **g,** Mean fluorescent intensity value of each droplet are plotted in the scatter plot, the positive droplets are the ones with a fluorescence intensity exceeding the threshold. For the final group comparison, all the droplet images from different groups are normalized before statistical calculation.

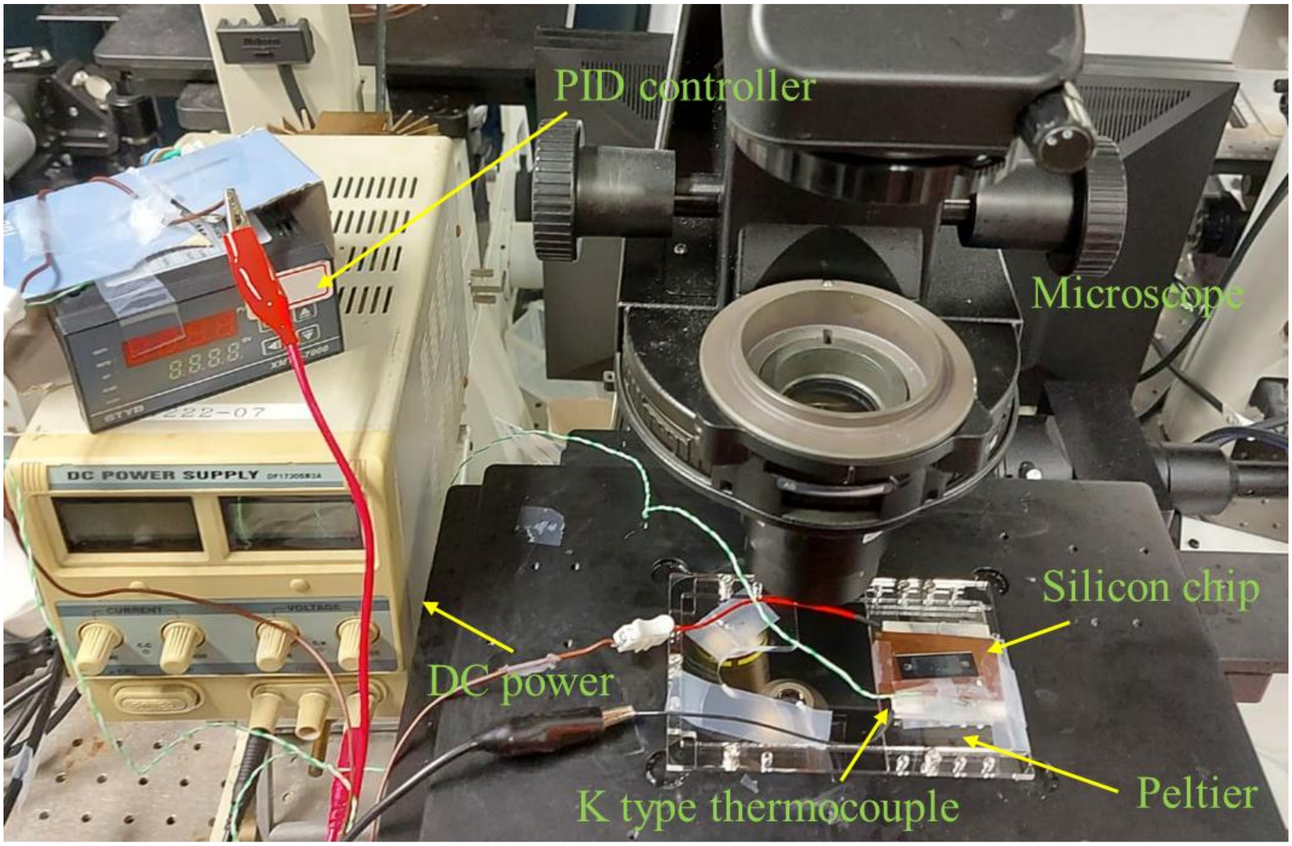

Figure S81. **The setup of real-time MEDICA**. The setup of the real-time MEDICA characterization contains a silicon observation chip, a Peltier-based heating system and fluorescence microscopy. After the sample loaded into the silicon chip and is sealed by qPCR film, the heating system is turned on to 37 $^{o}$C. then fluorescence image was recorded at the same location every min for 30 min.

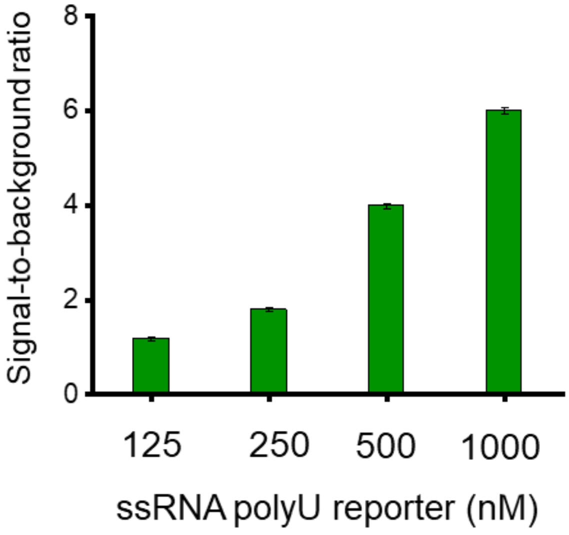

Figure S12. **The signal-to-background ratio versus the reporter concentration**. We tested a serial concentration of ssRNA reporter from 125 nM to 1000 nM following the MEDICA protocol. Each experiment was tested 3 times.

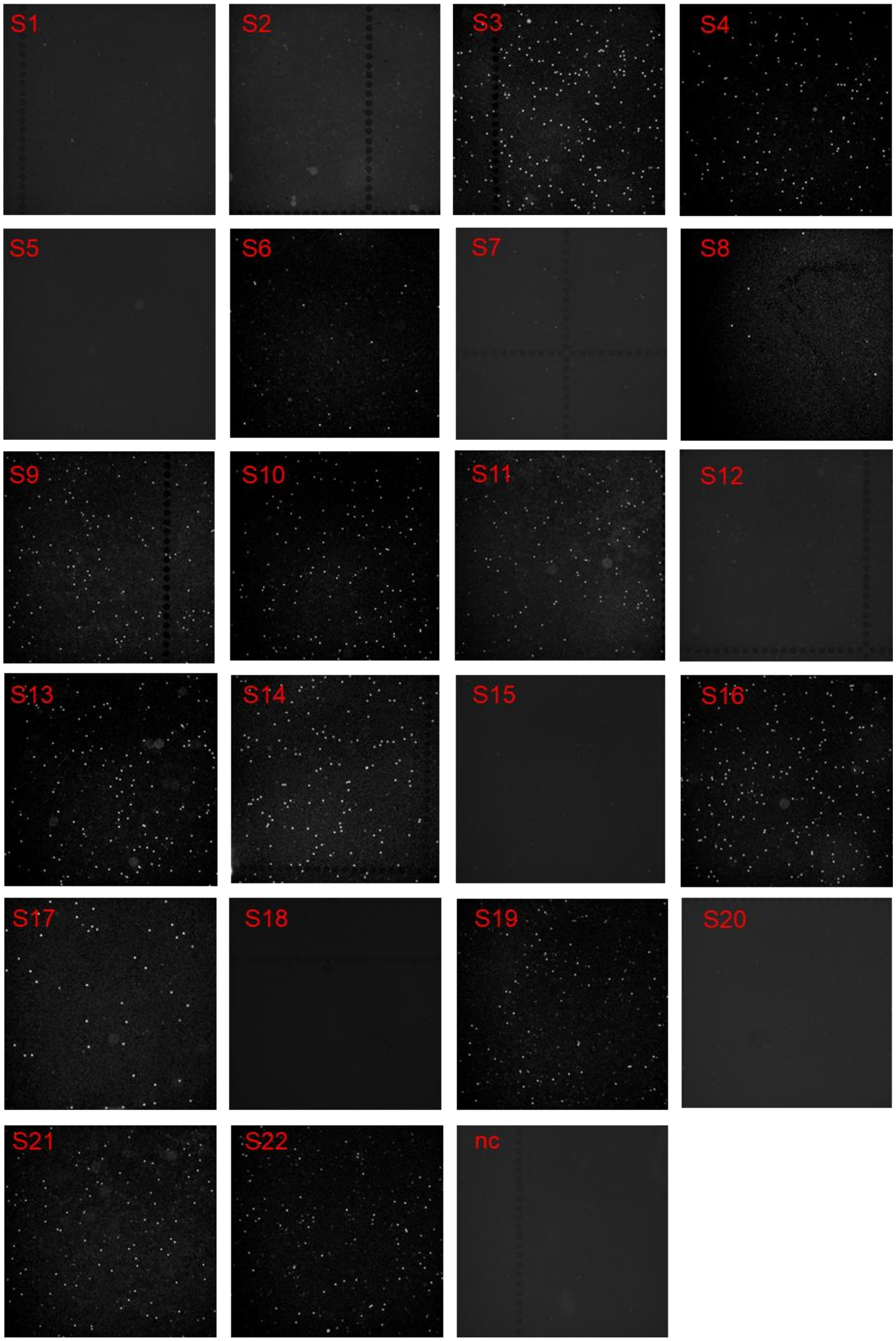

Figure S93. End-point fluorescence images of MEDICA for detection of HPV 16 using **22 clinical samples and NTC .**

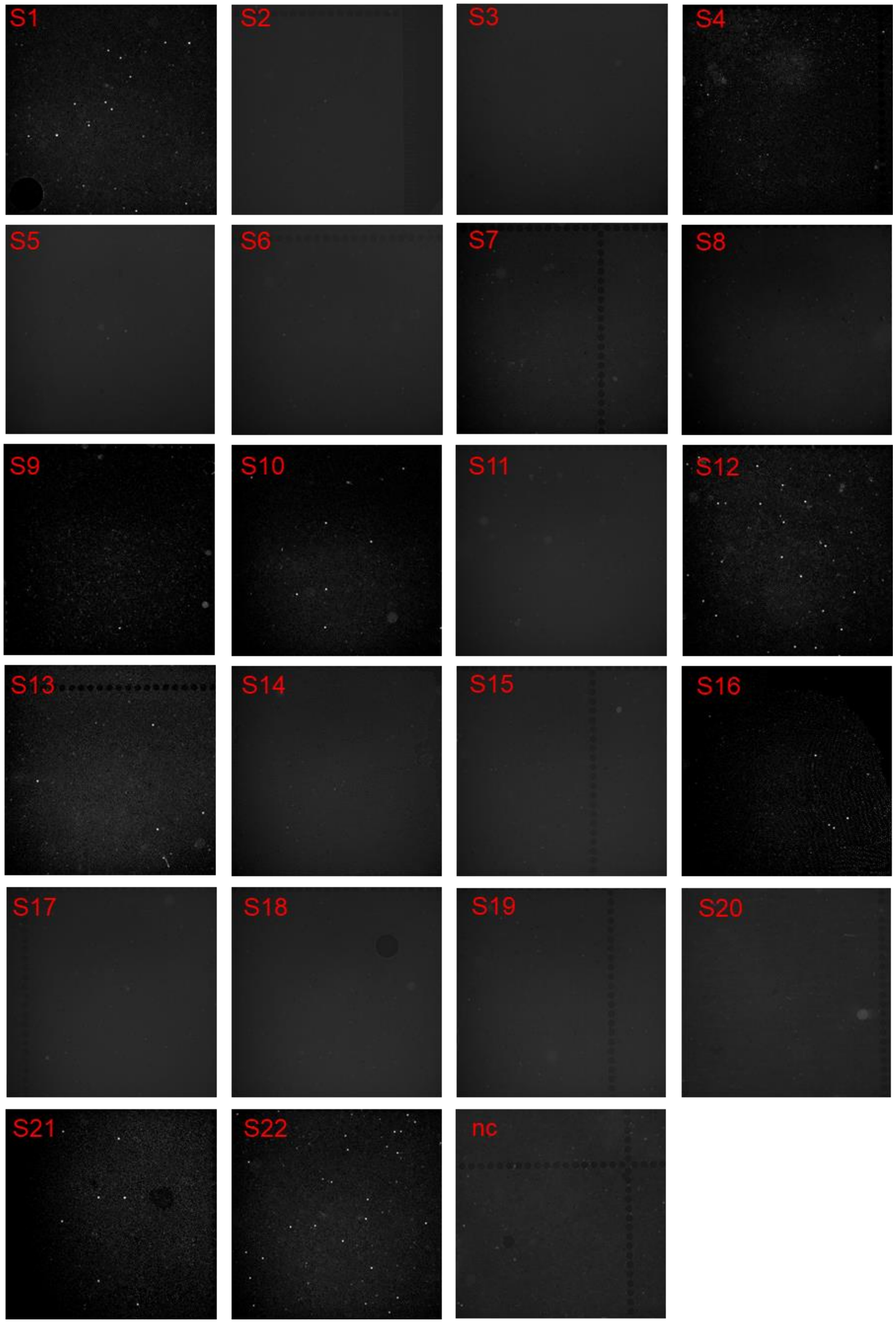

Figure S104. End-point fluorescence images of MEDICA for detection of HPV 18 using 22 clinical samples and NTC.

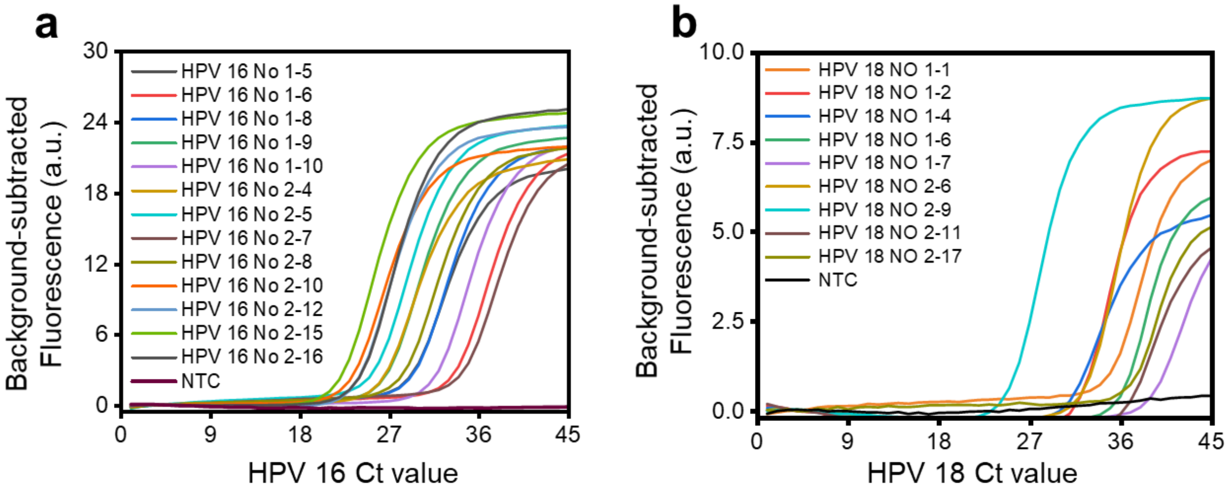

Figure S115. **qPCR results of clinical identification. a**, all HPV 16 positive patients and **b**, all HPV 18 positive patients.

Table 1. Comparison of MEDICA with other CRISPR nucleic acid detection methods

| Name | Enzyme | Preamplification | Steps | Reaction time (min) | Sample | Readout | quantitation | LOD (copies/mL) |
|---|---|---|---|---|---|---|---|---|
| DETECTR | LbCas12a | RPA | 2 | 60-120 | Crude extraction DNA | Fluorescence | No | 600 |
| HOLMESV2 | AsCas12a | LAMP | 1 or 2 | 120 | Synthetic DNA | Fluorescence | Qualitative | 6,000 (two step) |
| SHERLOCKv2 | LwaCas13a | RPA | 2 | 60-180 | Column-based DNA | Fluorescence or LFA | No | 4.8 |
| SHERLOCKv2 | LwaCas13a | RPA | 1 | 60-180 | Column-based DNA | Fluorescence or LFA | Qualitative | 12,000 |
| SHINE | LwaCas13a | RPA | 1 | 50 | Crude extraction RNA | Fluorescence or LFA | Qualitative | 5,000 |
| STOPCOVID | AspCas12b | LAMP | 1 | 60 | Crude extraction RNA | Fluorescence or LFA | Qualitative | 2,000 |
| DECOVID | LbCas12a | RPA | 1 | 15 – 30 | Synthetic RNA | Digital Fluorescence | Absolute | 1,000 |
| AIOD-CRISPR | LbCas12a | RPA | 1 | 40 | IVT RNA | Fluorescence | Qualitative | 180 |
| **MEDICA** | **LwaCas13a** | **RPA** | **1** | **10-25** | **Column-based DNA** | **Digital Fluorescence** | **Absolute** | **300** |

Table 1. The $C_t$ values of qPCR clinical tested results for enhanced one-pot reaction.

| sample No | Type | tested result | $C_t$ |
|---|---|---|---|
| B1 | DNA | HPV 16 | 22.3 |
| B2 | DNA | HPV 16 | 20.1 |
| B3 | DNA | HPV 16 | 21.9 |
| B4 | DNA | HPV 16 | 21.8 |
| B5 | DNA | NG | - |
| B6 | DNA | NG | - |
| B7 | DNA | CT | - |
| B8 | DNA | HPV 18 | 34.8 |
| B9 | DNA | HPV 18 | 31.7 |
| B10 | DNA | NG | - |
| B11 | DNA | NG | - |
| B12 | DNA | CT | - |

Table 2. The $C_t$ values of qPCR for detection of HPV16 using clinical samples

| sample No | Type | tested result | $C_t$ |
|---|---|---|---|
| 1 | DNA | HPV 18 | 32.3 |
| 2 | DNA | NA | - |
| 3 | DNA | NA | - |
| 4 | DNA | HPV 18 | 38.8 |
| 5 | DNA | NA | - |
| 6 | DNA | NA | - |
| 7 | DNA | NA | - |
| 8 | DNA | NA | - |
| 9 | DNA | HPV 18 | 36.7 |
| 10 | DNA | HPV 18 | 34.8 |
| 11 | DNA | NA | - |
| 12 | DNA | HPV 18 | 31.7 |
| 13 | DNA | HPV 18 | 35.4 |
| 14 | DNA | NA | - |
| 15 | DNA | NA | - |
| 16 | DNA | HPV 18 | 36.4 |

| | | | |
|---|---|---|---|
| 17 | DNA | NA | - |
| 18 | DNA | NA | - |
| 19 | DNA | NA | - |
| 20 | DNA | NA | - |
| 21 | DNA | HPV 18 | 34.7 |
| 22 | DNA | HPV 18 | 30.6 |

Table 3. The $C_t$ values of qPCR for detection of HPV18 using clinical samples

| sample No | Type | tested result | $C_t$ |
|---|---|---|---|
| 1 | DNA | NA | - |
| 2 | DNA | NA | - |
| 3 | DNA | HPV-16 | 21.9 |
| 4 | DNA | HPV-16 | 26.1 |
| 5 | DNA | NA | - |
| 6 | DNA | HPV-16 | 33.5 |
| 7 | DNA | NA | - |
| 8 | DNA | HPV-16 | 34.4 |
| 9 | DNA | HPV-16 | 28.2 |
| 10 | DNA | HPV-16 | 25.6 |
| 11 | DNA | HPV-16 | 25.4 |
| 12 | DNA | NA | - |
| 13 | DNA | HPV-16 | 23.6 |
| 14 | DNA | HPV-16 | 26.9 |
| 15 | DNA | NA | - |
| 16 | DNA | HPV-16 | 22.9 |

| | | | |
|---|---|---|---|
| 17 | DNA | HPV-16 | 31.4 |
| 18 | DNA | NA | - |
| 19 | DNA | HPV-16 | 29.2 |
| 20 | DNA | NA | - |
| 21 | DNA | HPV-16 | 26.5 |
| 22 | DNA | HPV-16 | 29.0 |

Table 4. Chemicals used in this work

| Reagent | Company | Experiment | Number |
| --- | --- | --- | --- |
| Benzonase nuclease | Sigma-Aldrich | Cas13a extraction | E1014-25KU |
| cOmplete Ultra Tablets, Mini, EDTA-free | Sigma-Aldrich | Cas13a extraction | 05892791001 |
| Lysozyme from chicken egg white | Sigma-Aldrich | Cas13a extraction | 7374252 |
| Strep-Tactin Superflow Plus resin | Qiagen | Cas13a extraction | 30004 |
| SUMO protease | Thermo Fisher Scientific | Cas13a extraction | 12588018 |
| NP-40 Surfact-Amps Detergent Solution | Thermo Fisher Scientific | Cas13a extraction | 85124 |
| TwistAmp Basic | TwistDx | MEDICA | TABAS03KIT |

| | | | |
|---|---|---|---|
| Trizma® base | Sigma-Aldrich | Buffer optimization | 93352-500G |
| HEPES | Sigma-Aldrich | Buffer optimization | H4034-1KG |
| Sodium chloride | Sigma-Aldrich | Buffer optimization | S9888-1KG |
| Potassium chloride | Sigma-Aldrich | Buffer optimization | P4504-500G |
| Ammonium sulfate | Sigma-Aldrich | Buffer optimization | A4418-500G |
| Potassium acetate | Sigma-Aldrich | Buffer optimization | P1147-500G |
| Poly(ethylene glycol) | Sigma-Aldrich | Buffer optimization | P2139-500G |
| Bovine Serum Albumin | Sigma-Aldrich | Buffer optimization | A9418-10G |
| DL-Dithiothreitol solution | Sigma-Aldrich | Buffer optimization | 43816-50ML |

| | | | |
|---|---|---|---|
| TWEEN® 20 | Sigma-Aldrich | Buffer optimization | P9416-50ML |
| SU8 | HKUST NFF | Chip fabrication | |
| 3M™ Novec™ 7500 | 3M | Droplet partition | |
| EA surfactant | Thunderbio | Droplet partition | |
| rNTP Mix | New England Biolabs | MEDICA | N0466L |
| RNase Inhibitor, Murine | New England Biolabs | MEIDCA | M0314L |
| NxGen® T7 RNA Polymerase | Lucigen | MEDICA | 30223-1 |
| Ex Taq DNA polymerase | TAKARA | qPCR | R028A |
| Qiagen kit | Qiagen | Clinical extraction | 51304 |

Table 5. Sequences used in this work

| Reagent | Sequences (from 5 to 3) | Company | notes |
|---|---|---|---|
| HPV 16 L1 gene | GGTCTACTGCAAATTTAGCCAGTTCAAA TTATTTTCCTACACCTAGTGGTTCTATGG TTACCTCTGATGCCCAAATATTCAATAA ACCTTATTGGTTACAACGAGCACAGGGC CACAATAATGGCATTTGTTGGGGTAACC AACTATTTGTTACTGTTGTTGATACTACA CGCAGTACAAATATGTCATTATGTGCTG CCATATCTACTTCAGAAACTACATATAA AAATACTAACTTTAAGGAGTACCTACGA CATGGGGAGGAATATGATTTACAGTTTA TTTTTCAACTGTGCAAAATAACCTTAACT GCAGACGTTATGACATACATACATTCTA TGAATTCCACTATTTTGGAGGACTGGAA TTTTGGTCTACAACCTCC | genscript | Cloning Vector: pUC57 |

| | | | |
|---|---|---|---|
| HPV 18<br>E1 gene | ATGGCTGATCCAGAAGGTACAGACGGGG<br>AGGGCACGGGTTGTAACGGCTGGTTTTA<br>TGTACAAGCTATTGTAGACAAAAAAACA<br>GGAGATGTAATATCTGATGACGAGGACG<br>AAAATGCAACAGACACAGGGTCGGATAT<br>GGTAGATTTTATTGATACACAAGGAACA<br>TTTTGTGAACAGGCAGAGCTAGAGACAG<br>CACAGGCATTGTTCCATGCGCAGGAGGT<br>CCACAATGATGCACAAGTGTTGCATGTT<br>TTAAAACGAAAGTTTGCAGGAGGCAGCA<br>AAGAAAACAGTCCATTAGGGGAGCGGCT<br>GGAGGTGGATACAGAGTTAAGTCCACGG<br>TTACAAGAAATATCTTTAAATAGTGGGC<br>AGAAAAAG | genscript | Cloning<br>Vector:<br>pUC57 |
| Coronaviru<br>s ORF ab | GGGCATTGATTTAGATGAGTGGAGTATG<br>GCTACATACTACTTATTTGATGAGTCTGG<br>TGAGTTTAAATTGGCTTCACATATGTATT<br>GTTCTTTCTACCCTCCAGATGAGGATGA<br>AGAAGAAGGTGATTGTGAAGAAGAAGA | genscript | Cloning<br>Vector:<br>pUC57 |

| | | | |
|---|---|---|---|
| | GTTTGAGCCATCAACTCAATATGAGTAT<br>GGTACTGAAGATGATTACCAAGGTAAAC<br>CTTTGGAATTTGGTGCCACTTCTGCTGCT<br>CTTCAACCTGAAGAAGAGCAAGAAGAA<br>GATTGGTTAGATGATGATAGTCAACAAA<br>CTGTTGGTCAACAAGACGGCAGTGAGGA<br>CAATCAGACAACTACTATTCAAACAATT<br>GTTGAGGTTCAACCTCAATTAGAGATGG<br>AACTTACACCAGTTGTTCAGACTATTGA<br>AGTGAATAGTTTTAGTGGTTATTTAAAA<br>CTTACTGACAAT | | |
| Coronaviru<br>s N | CAAAAGGCTTCTACGCAGAAGGGAGCA<br>GAGGCGGCAGTCAAGCCTCTTCTCGTTC<br>CTCATCACGTAGTCGCAACAGTTCAAGA<br>AATTCAACTCCAGGCAGCAGTAGGGGAA<br>CTTCTCCTGCTAGAATGGCTGGCAATGG<br>CGGTGATGCTGCTCTTGCTTTGCTGCTGC<br>TTGACAGATTGAACCAGCTTGAGAGCAA<br>AATGTCTGGTAAAGGCCAACAACAACAA | genscript | Cloning<br>Vector:<br>pUC57 |

| | | |
|---|---|---|
| | GGCCAAACTGTCACTAAGAAATCTGCTG<br>CTGAGGCTTCTAAGAAGCCTCGGCAAAA<br>ACGTACTGCCACTAAAGCATACAATGTA<br>ACACAAGCTTTCGGCAGACGTGGTCCAG<br>AACAAACCCAAGGAAATTTTGGGGACCA<br>GGAACTAATCAGACAAGGAACTGATTAC<br>AAACATTGGCCGCAAATTGCACAATTTG<br>CCCCCAGCGCTTCAGCGTTCTTCGGAAT<br>GTCGCGCATTGGCATGGAAGTCACACCT<br>TCGGGAACGTGGTTGACC | |
| HPV16-<br>L1-13-<br>RPA-<br>Forward | GAAATTAATACGACTCACTATAGGGTTG<br>TTGGGGTAACCAACTATTTGTTACTGTT | IDT |
| HPV16-<br>L1-13-<br>RPA-<br>Reverse1 | CCTCCCCATGTCGTAGGTACTCCTTAAAG | IDT |
| HPV18-<br>E1-13- | GAAATTAATACGACTCACTATAGGGTCG<br>GATATGGTAGATTTTATTGATACACA | IDT |

| | | |
|---|---|---|
| RPA-Forward1 | | |
| HPV18-E1-13-RPA-Reverse1 | CATGCAACACTTGTGCATCATTGTGGACCT | IDT |
| Orf1ab-cas13a-RPA-Forward | GAAATTAATACGACTCACTATAGGGCCAAGGTAAACCTTTGGAATTTGGTGCCAC | IDT |
| Orf1ab-cas13a-RPA-Reverse | ACTATCATCATCTAACCAATCTTCTTCTTG | IDT |
| Ngene-cas13a-RPA-Forward | GAAATTAATACGACTCACTATAGGGCGGCAGTCAAGCCTCTTCTCGTTCCTCATC | IDT |
| Ngene-cas13a- | CAGACATTTTGCTCTCAAGCTGGTTCAATC | IDT |

| | | |
|---|---|---|
| RPA-Reverse | | |
| HPV16-E7-13-PCR-Forward | AGCTCAGAGGAGGAGGATGAA | IDT |
| HPV16-E7-13-PCR-Reverse | GGTTACAATATTGTAATGGGCTC | IDT |
| HPV18-E1-PCR-Forward | CATTTTGTGAACAGGCAGAGC | IDT |
| HPV18-E1-PCR-Reverse | ACTTGTGCATCATTGTGGACC | IDT |
| ZIKA Forward | GAAATTAATACGACTCACTATAGGGCGGAACTCCACACTGGAACAACAAA | IDT |
| ZIKA Reverse | TGCACCATCCATCTCAGCCTCCAGAGCTCC | IDT |

| | | |
|---|---|---|
| ORF1ab-Cas13a-crRNA | GAUUUAGACUACCCCAAAAACGAAGGGGACUAAAACCUCUUCUUCAGGUUGAAGAGCAGCAGAA | Synthego |
| Ngene-cas13a-crRNA | GAUUUAGACUACCCCAAAAACGAAGGGGACUAAAACGCAAAGCAAGAGCAGCAUCACCGCCAUU | Synthego |
| HPV16-L1-Cas13a-crRNA | GAUUUAGACUACCCCAAAAACGAAGGGGACUAAAACUCUGAAGUAGAUAUGGCAGCACAUAAUG | Synthego |
| HPV18-E1-Cas13a-crRNA | GAUUUAGACUACCCCAAAAACGAAGGGGACUAAAACUGCUGUCUCUAGCUCUGCCUGUUCACAA | Synthego |
| ZIKA crRNA | GAUUUAGACUACCCCAAAAACGAAGGGGACUAAAACACUCCCUAGAACCACGACAGUUUGCCUU | Sangon |
| HPV18E1-PCR | /56-FAM/AGAGACAGCACAGGCATTGTTCCATG/36-TAMSp/ | IDT |

| | | |
|---|---|---|
| HPV16L1-PCR*(6)* | /56-FAM/GTCATTATGTGCTGCCATATCTACTTC/36-TAMSp/ | IDT |
| PolyU reporter | /56-FAM/rUrUrUrUrU/3IABkFQ/ | IDT |
